# Estimating the effects of reduced sunlight due to solar geoengineering on suicide in the United States

**DOI:** 10.1101/2022.10.08.22280867

**Authors:** Shinsuke Tanaka, Tetsuya Matsubayashi

**Affiliations:** Tufts University. 160 Packard Ave. Medford, MA 02155, USA; Osaka University. 1-31 Machikaneyama Toyonaka, Osaka, 560-0043 Japan

## Abstract

**Background:** Solar geoengineering, whereby sunlight is reflected back into space at the outer atmosphere to reduce incoming sunlight, is increasingly considered a viable option to mitigate global warming, yet the health consequences of reducing incoming sunlight remain poorly quantified.

**Objectives:** This study examines the effects of sunlight exposure on the rate of suicide across the United States over nearly three decades and projects the impact of geoengineering-induced reductions in sunlight on suicides by 2100.

**Methods:** The analysis relates sunlight exposure, as measured by solar insolation, to the suicide rate at the county-by-month level in the United States between 1979 and 2004 (N = 444,861), after adjusting for temperature, precipitation, county-by-month effects, and state-by-year effects. We project the excess suicides due to the negative radiative forcing required to keep the temperature rise below 1.5 °C by 2100.

**Results:** We find that suicide rates increase by 6.99% (95% CI: 3.86, 10.13) as sunlight decreases by one standard deviation, which is almost equivalent to the difference in sunlight between the lowest (Vermont) and highest (Arizona) state-level averages. The effects are similar across an extensive set of county characteristics and over time, suggesting limited adaptation to sunlight exposure in suicidal behavior. We also find that insufficient sunlight exposure increases the searches containing depressive language on Google Trends. These estimates suggest that solar geoengineering could result in 1.26–3.18 thousand additional suicides by 2100 under the business-as-usual scenario, which could more than offset the suicides averted by temperature fall.

**Discussion:** Our findings highlight the substantial benefits of sunlight exposure on the incidence of suicide and mental well-being, thus calling for climate policy to better balance the potential benefits and harms of solar geoengineering.

## Introduction

Climate change threatens human health and well-being in numerous dimensions. Despite the worldwide efforts to curb the temperature rise and combat climate change, culminating in the Paris Agreement, progress in reducing pollution emissions is insufficient, and key mechanisms to accomplish the goal of keeping global warming to less than 1.5 °C remain to be developed. In this light, there is increasing interest in employing new climate geoengineering technologies. One such technology is solar radiation management, which reflects sunlight back into space by using orbiting mirrors or spraying aerosol particles into the outer atmosphere (1–3). While the low financial cost and high availability of relevant technologies mean that solar geoengineering could be implemented within a couple of years, resulting in a rapid reduction in global temperatures (4), the substantial uncertainties that remain concerning the impacts of solar geoengineering on both the natural environment and human health and well-being have given rise to controversies among scholars and policymakers regarding its deployment (5–11).

This study is, to the best of our knowledge, one of the first attempts to consider the potential consequences of proposed solar geoengineering for human well-being. Specifically, we examine how the reduction of incoming sunlight through solar geoengineering may affect our mental health, focusing on suicide as an outcome. Our analysis involves two steps. We first quantify the effects of solar insolation on suicide rates using novel datasets at the county-by-month level between 1979 and 2004 from the United States (N = 444,861). We then project the impacts of proposed solar geoengineering to accomplish the goal of keeping the temperature rise below 1.5 °C by 2100 by assessing the excess suicides due to the negative radiative forcing relative to the averted suicides due to reduced temperatures.

The linkage between sunlight and human health naturally varies depending on outcomes to be studied. On the one hand, limiting exposure to sunlight can be beneficial because the Sun’s ultraviolet (UV) rays cause skin cancer. The growing awareness of the adverse effects of UV rays, amplified by public campaigns to reduce excessive exposure to sunlight (12), has resulted in people spending increasing amounts of time indoors. On the other hand, there is increasing evidence that insufficient sunlight exposure can also be hazardous. For example, the worldwide vitamin D deficiency (about 40% of population in the U.S. and Europe (13,14)) due to increased time indoors has been linked to increased mortality from chronic diseases such as cancer, cardiovascular diseases, and metabolic syndrome, resulting in an estimated 340,000 deaths in the U.S. and 480,000 deaths in Europe each year (15).

The potential effects of sunlight exposure on mental health, and on suicide in particular, remain surprisingly poorly understood. Insufficient sunlight exposure during wintertime has been hypothesized to explain Seasonal Affective Disorder (SAD), a type of depression that peaks in wintertime, because of disrupted sleep, impeded neurotransmissions of serotonin, vitamin D deficiency, and the overproduction of melatonin (16–18). In contrast, suicide rates typically peak in late spring to early summer and decline in wintertime (Figure S1, 19). This apparently paradoxical seasonal pattern in the suicide rate has been cited as discrediting the sunlight-suicide relationship.

Such disparities in evidence merit close attention because suicide is one of the major causes of death, especially among youth worldwide. Moreover, those investigating the so-called “Deaths of Despair” (20) phenomenon attribute the rising rate of premature deaths and declining life expectancy among certain demographic groups in the U.S to an increase in the suicide rate. Since suicide has most often been linked to individual, social, and economic factors (21), policy and program interventions have tended to address the issue accordingly. Yet, by and large these efforts have failed to stem the recent rising trend in the rates of suicide—by 30% between 2000 and 2018 and more than twice higher among younger people (22)— which has imposed a major public health burden in critical need of a new approach. New research is calling into question whether environmental changes, e.g., temperature (23,24) and air pollution (25), portend a substantial change in the suicide rate.

Our empirical approach involves two novel features. First, sunlight exposure is measured by solar insolation, which is the amount of incident solar radiation energy received from the Sun per unit surface of the Earth over a specified period. Many factors determine how much sunlight reaches the surface, such as the solar zenith angle, the variable distance from the Earth to the Sun, day length, weather conditions, levels of atmospheric aerosols, and solar activity. Thus, solar insolation measures the intensive margin of sunlight exposure to humans much more precisely than the duration of daylight, the metric most widely used in the existing literature. Mixed evidence in the literature may arise from the conventional use of daylight duration that measures only the extensive margin of sunrise and sunset and allows for little variation after controlling for the month effects (26–31). Further, the more direct measure of sunlight exposure provided by solar insolation is necessary to project the impacts of geoengineering-driven reductions in sunlight on suicides.

Second, our longitudinal dataset with substantial cross-sectional and temporal coverage, along with our novel use of a direct measure of solar insolation, offers a unique opportunity to plausibly isolate the effects of sunlight exposure on suicide from local monthly seasonality effects, such as the daylight duration and the school calendar, as well as local time-varying shocks, such as economic conditions, agricultural production, poverty rates, and gun ownership (Methods). In contrast, a handful of studies using solar insolation or irradiation as the exposure measure have relied exclusively on time-series data and showed a positive association with suicide (30), which is likely to be driven by other local seasonal patterns in meteorological, social, and economic factors.

Our analysis relates the county-monthly suicide rate to solar insolation during that month in that county, after adjusting for local temperature and precipitation and nonparametrically controlling for county-by-month and state-by-year effects. The basic intuition behind this model is that the effects of solar insolation are estimated based on the random variation in the amount of sunlight between, for example, August 2000 and August 2001 in the same county. The model assumption is that such within-county variation in the amount of sunlight for a given month across years is uncorrelated with any other factors that affect the suicide rate. Such assumption is plausible because the amount of sunlight is determined by random fluctuations in climatic and meteorological conditions, after adjusting for factors related to sunlight itself, such as temperature and precipitation. Thus, our estimates are not confounded by the permanent heterogeneities across counties or state-specific within-year fluctuations in sunlight and suicide rate. For example, individuals with higher incomes may be at a lower risk of suicides and may be inclined to live in areas with sunnier climates; or state-level annual average suicide rates may be lower in years with greater sunlight and greater agricultural production.

The main analysis considers several lagged impacts of sunlight exposure by estimating distributed-lag models that allow for the cumulative, or displacement, effects of sunlight exposure in given and past months on the suicide rate in the given month. As a placebo test, we also include sunlight in the following month to confirm that future sunlight exposure is unrelated to the suicide rate in a given month. While the main analysis focuses on the level-log model, we also allow for nonlinear impacts of sunlight by adopting a high-order polynomial function and a nonparametric approach as robustness checks (Method).

We present several additional analyses. First, we apply a comparable model to investigate the potential heterogeneities in the effects of sunlight by stratifying the sample based on county-level averages of temperature, income, air conditioner ownership, and gun ownership. We also explore impacts by gender with different suicide methods, e.g., violent vs. non-violent. Second, we investigate the impacts of sunlight in each year over our study period to look for potential adaptation. Given that increasing public awareness of the harmful effects of sunlight exposure (e.g. skin cancer) has caused people increasingly to avoid such exposure, we would expect to see the suicide effects of sunlight lessen in more recent years. Third, to uncover potential mechanisms, we assess whether sunlight exposure affects mental well-being among a general population by exploring whether the number of searches containing or related to depressive language (Methods) on Google is related to patterns of solar insolation.

Finally, we project the first estimates, to the best of our knowledge, of the impacts of solar radiation management geoengineering on suicides by 2100. We consider the negative radiative forcing gap required to keep the temperature rise below 1.5 °C, as targeted by the Paris Agreement, from the business-as-usual emissions scenario in each year. We then compute the excess suicides induced by reduced sunlight. Given that reduced temperature has been shown to reduce suicides (24), we incorporate suicides averted by temperature fall to arrive at the net impacts of solar geoengineering on the incidence of suicides.

## The effects of sunlight on the suicide rate

We find that insufficient sunlight significantly increases the suicide rate. The results of the distributed lag model show that the contemporaneous effect is negative and statistically significant (*β*_0_ =−0.049; 95% Confidence Interval (CI): −0.092, −0.006) (Table S2). We additionally find a statistically significant and larger in magnitude effect of sunlight from the previous month (*β*_−1_ =−0.085; 95% CI: −0.132, −0.038), suggesting that the sunlight-suicide relationship is a dynamic one in which the trajectory of depression leading to suicide can develop over months (32). In contrast, the effect of sunlight in the second previous month is negligible (*β*_−2_ =−0.003; 95% CI: −0.046, 0.040), indicating that the cumulative effects span two months. As placebo evidence, we find that the effect of sunlight in the following month has no impact on the suicide rate in this month (*β*_1_ =0.010; 95% CI: −0.037, 0.057). The overall impact of sunlight is thus given by *β*_0_ + *β*_−1_, which is −0.134 (95% CI: −0.194, −0.074, *p*-value = 0.000, N = 444,861).

These estimates suggest that a one standard deviation decrease in population-weighted sunlight, 6,449.2 KJ/m^2^, from the population-weighted mean value of 16,422.9 KJ/m^2^, leads to a 6.99% (95% CI: 3.86, 10.13) increase in the suicide rate. The estimated size of the effect is nearly comparable to the effect of a one standard deviation increase in temperature on the suicide rate (Table S2).

We test the robustness of these estimates in several ways. First, we use different, and often more granular, fixed effects, such as county-by-month and county-by-year fixed effects, and cubic polynomial models. Figure 1 illustrates the estimated cumulative effects based on these alternative models. The shaded area represents the 95% CI of the baseline model with county-by-month and state-by-year fixed effects. We find that all estimated effects from the alternative models are quantitatively similar to each other. Second, we find similar results using various other dependent variables and a count model (Table S3). Third, we explore temporal displacement effects over a longer period, showing consistent evidence that the months before the second previous month have no impact on the suicide rate in a given month (Figure S4).

**Figure 1:**
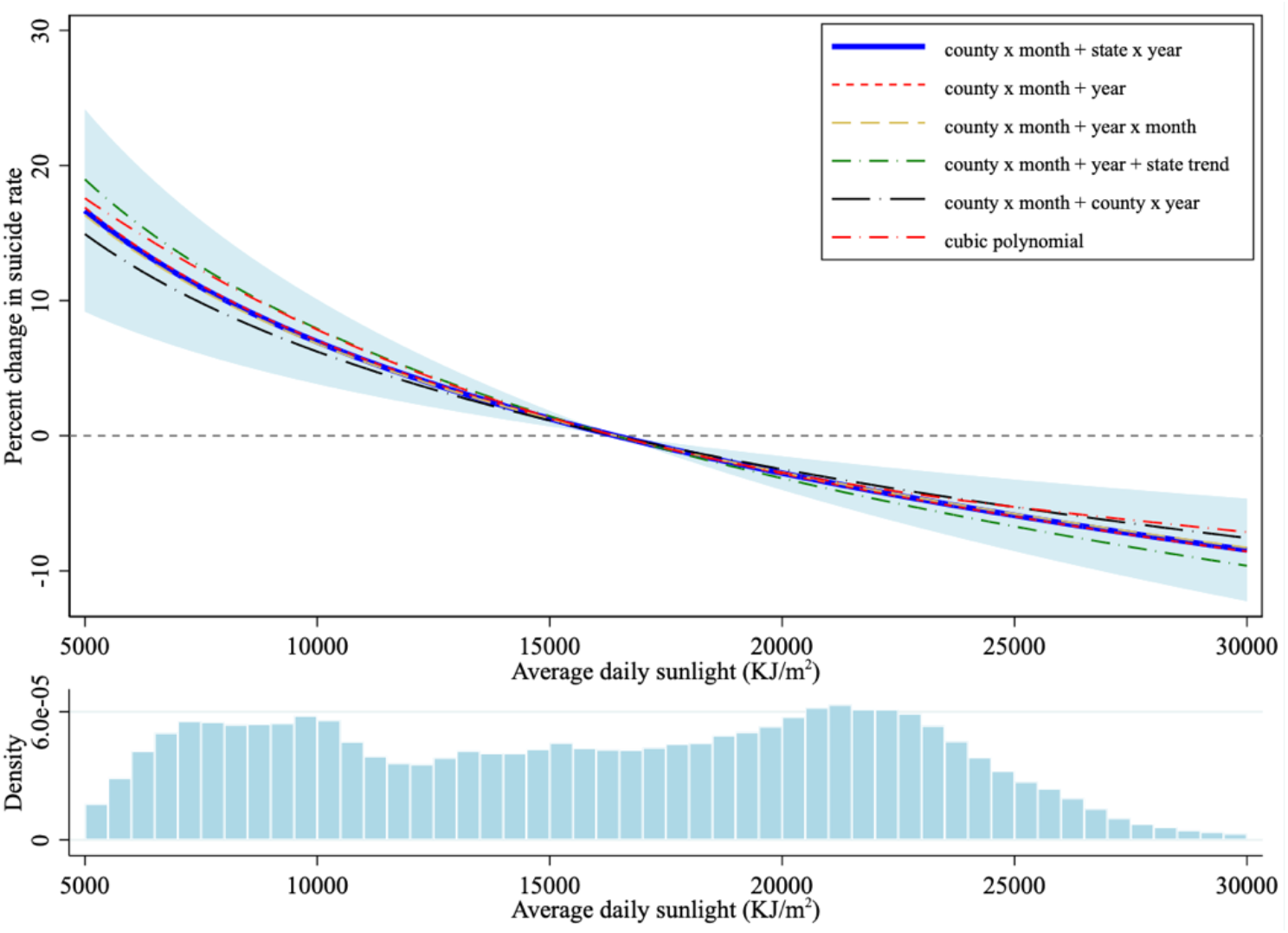
Effects of sunlight on the suicide rate *Notes*: The top panel plots the estimated cumulative effects of sunlight in given and previous months on the percentage change in the suicide rate with various time effects and models as specified by the labels. The intercepts are adjusted to allow all curves to show no change in the suicide rate at the mean sunlight level. The blue shaded area represents the 95% confidence interval of the elasticity estimated from the baseline model with the county-month and state-year fixed effects. All regressions additionally include the mean temperature and precipitation in the given and previous months. The underlying coefficients are presented in Table S2. The bottom figure plots the distribution of average daily sunlight in our sample. The average monthly suicide rate weighted by county population is 0.955 per 100,000 population. The mean of average daily sunlight weighted by county population is 16,422.91 KJ/m^2^.

### Heterogeneities in the effects of sunlight

We now explore potential heterogeneities in the sunlight-suicide relationships by county characteristics (Figure 2). To do so, we first compute the population-weighted county-level median characteristics, stratify the sample into the above- and below-median groups, and simultaneously estimate the coefficients for both groups (Methods). We find that the effects of sunlight are virtually identical between counties with above- or below-median sunlight, e.g., high-vs. low-latitude regions. The results from the nonparametric approach that explores the effects in each decile of historical sunlight exposure provide consistent results, although the highest decile appears to have a slightly stronger effect (Figure S2). We also find that the effects in each month are quantitatively in the similar range (Figure S3). These results suggest that the effects of sunlight exposure do not vary by the baseline levels of sunlight exposure. We also find similar impacts of sunlight on the suicide rate across county temperature, indicating that the effect of sunlight on the suicide rate is independent from that of temperature. It is worth noting that while the log of sunlight is positively correlated with the monthly temperature, temperature is significantly and positively associated with the suicide rate. Using the county-level income data, we find a larger effect of sunlight in counties with above-median income, whereas the estimated effect among below-median income counties is not statistically different from zero. We also find no heterogeneities in the effects of sunlight by the state-level adoption rate of air conditioning, although people with air conditioning may stay indoors on warm days and avoid sunlight exposure. Further, we find no heterogeneities by state-level gun ownership rate, although over 50% of suicides involved a firearm in 2020 (33). Lastly, we find no heterogeneities by method of suicide among men, whereas the estimated effects are smaller for women, particularly using violent methods.

**Figure 2:**
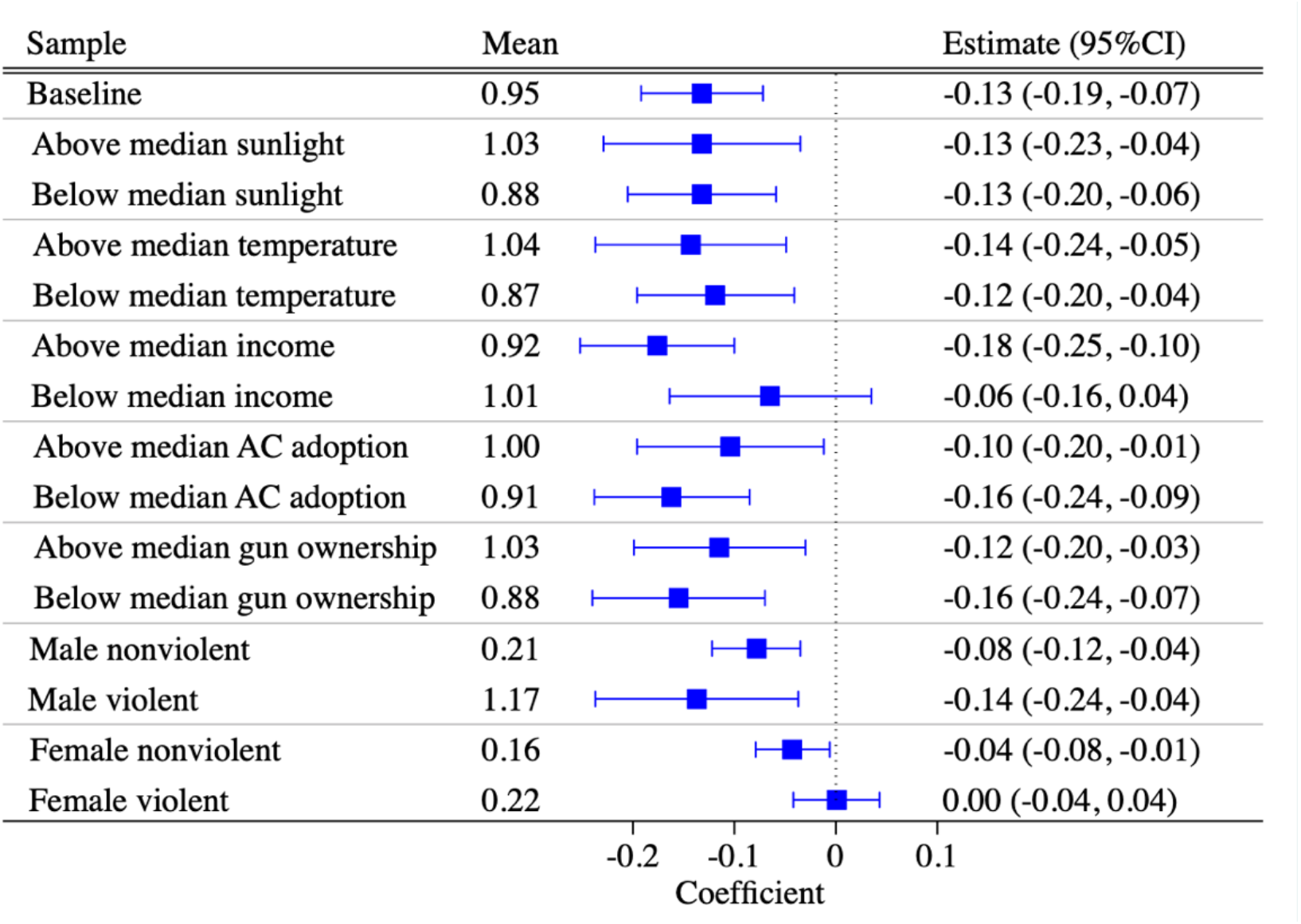
Heterogeneous effects of sunlight. *Notes*: This figure plots the cumulative effect of sunlight in given and previous months, as given by *β*_0_ + *β*_−1_ and its 95% confidence interval based on Equation (2) in Methods.

### Adaptation over time

Existing studies find mixed evidence regarding the extent of adaptation to changes in the environment. For example, while the effects of temperature on all-cause mortality have lessened over time in the U.S. due to the adoption of air conditioning (34), the effects of temperature on suicide have been stable over decades in the U.S. and Mexico (24).

In contrast, existing evidence offers no insight into the sunlight-suicide relationship. Increasing evidence regarding the role of ultraviolet radiation in skin cancer has led to reduced time spent outdoors over the past decades, which has potentially reduced the effects of sunlight on suicide over time. We find that the effects of sunlight on the suicide rate have been quantitively similar over our study period (Figure 3). Overall, these findings point to limited adaptation to sunlight exposure in suicidal behavior.

**Figure 3:**
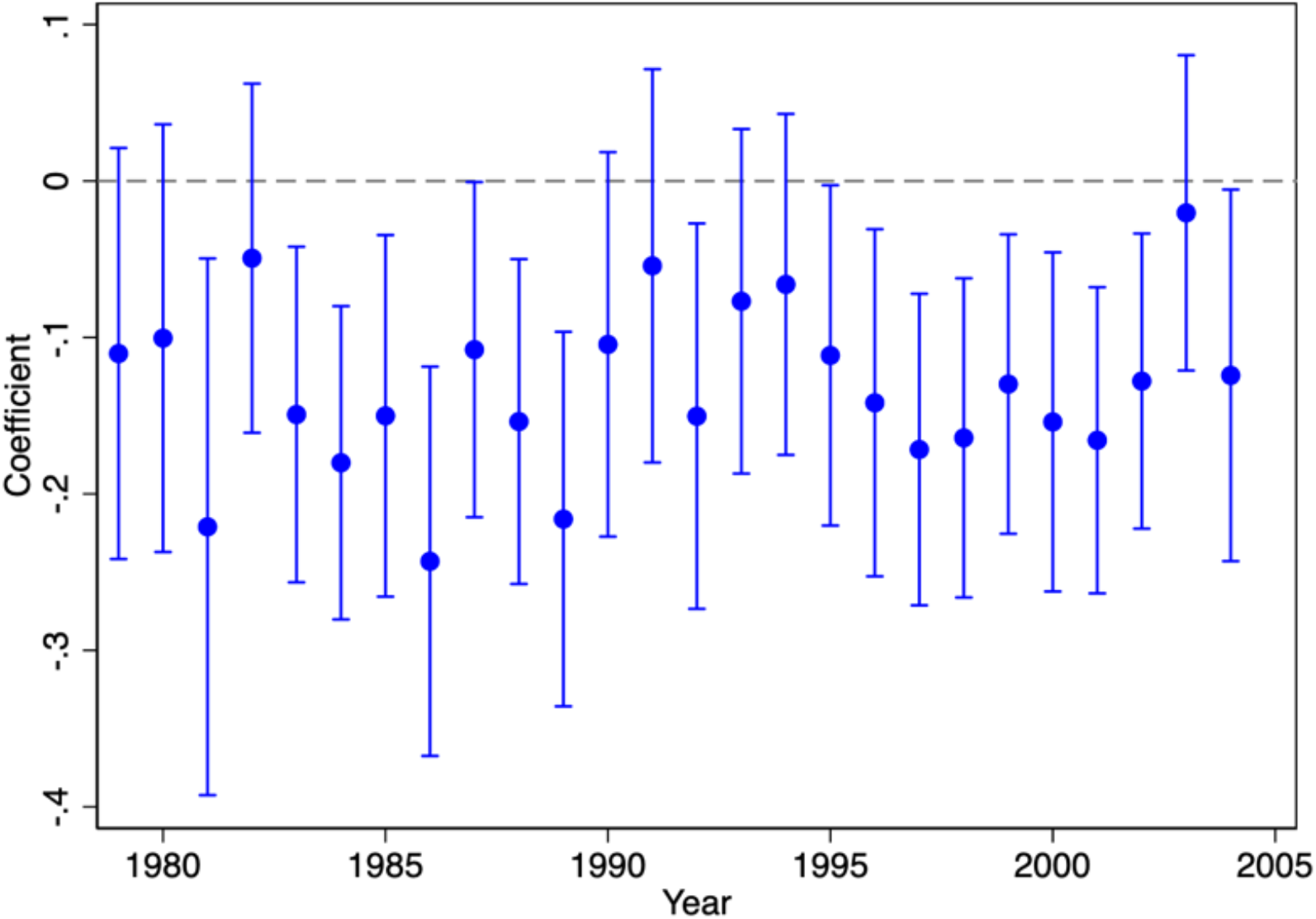
Effects of sunlight on suicide rate across years. *Notes*: This figure plots the cumulative effect of sunlight in given and previous months, as given by *β*_0_ + *β*_−1_ and its 95% confidence interval based on Equation (3) in Methods.

### Sunlight and depressive language in Google Trends

Our findings thus far demonstrate a strong relationship between sunlight and suicide, while the underlying mechanism remains unclear. We seek to uncover the mechanism by examining the possibility that sunlight exposure is related to mental well-being. Building upon Burke et al. (24), we measure mental well-being by the patterns of internet searches for or using depressive language (listed in Methods) among the general population, in anticipation that they vary in accordance with sunlight. To test this, we obtain the monthly internet search results using depressive language on Google using Google Trends between 2004 and 2011 (Methods), bounded by the initial year when Google Trends is available and the last year for which the sunlight data is available.

Using a similar fixed-effects model (Methods), we find evidence consistent with the main analysis that sunlight in given and previous months has significantly negative impacts on the number of searches employing depressive language (Figure 4). In particular, we find that a one standard deviation decrease in the amount of monthly sunlight (6,532.7 KJ/m^2^) at the state level leads to a 4.33% (95% CI: 3.26, 5.40, *p* = 0.000, N = 4,655) increase in the search volumes for depressive language. We also find similar effects with the Designated Marketing Area (DMA)-level of observations with various fixed effects (Figure 4). We further find comparable effects with subsets of the entire list of depressive language (Table S9).

**Figure 4:**
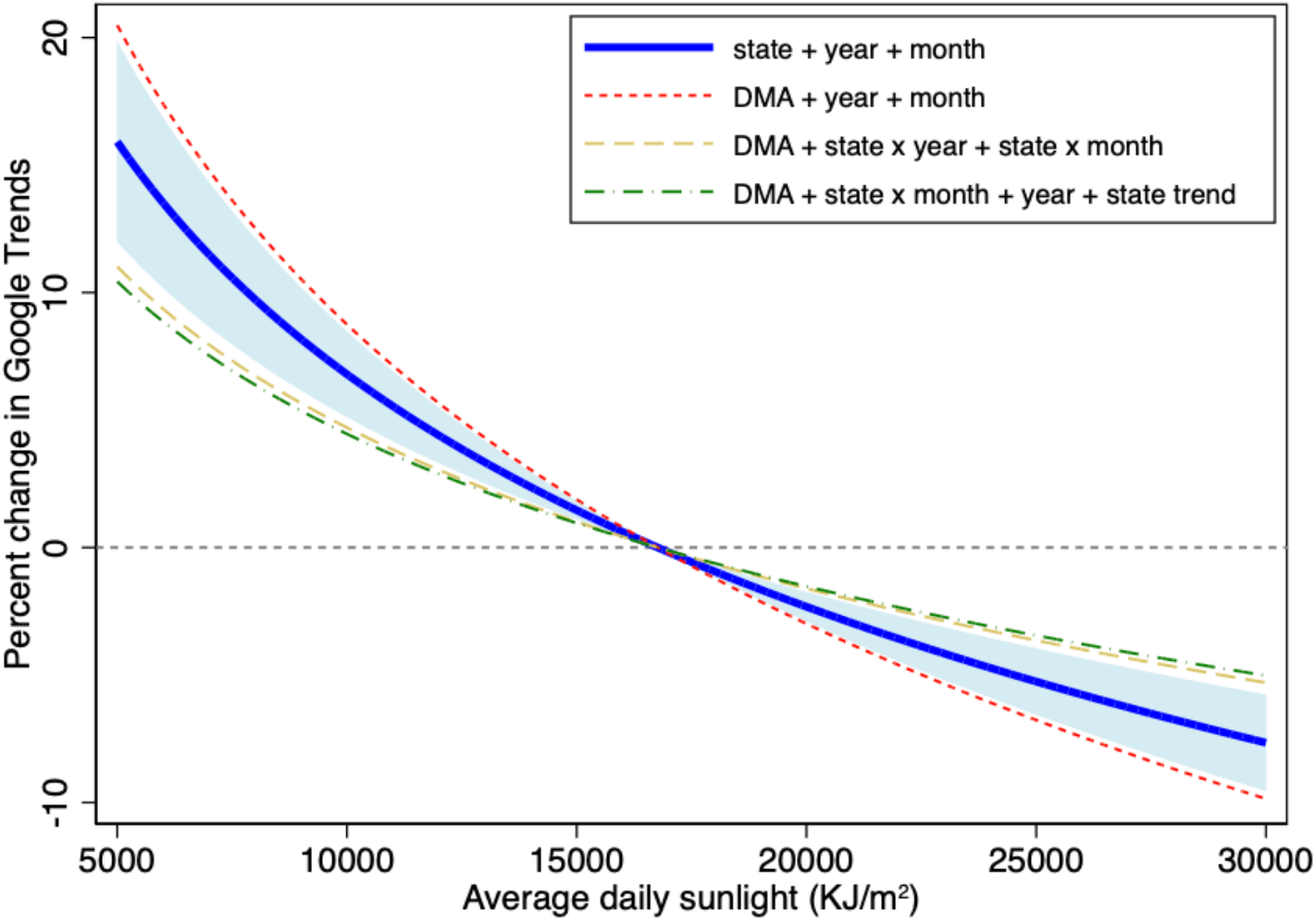
Effects of sunlight on depressive language searches from Google Trends. *Notes*: This figure plots the estimated cumulative effects of sunlight in given and previous months on the percentage change in Google Trends search interests for the set of depressive language terms as specified in Methods with various time effects as specified by the labels. The intercepts are adjusted to allow all curves to show no change in Google Trends values at the mean sunlight level. The thick blue line indicates the estimate from the observations at the state-year-month level, and the blue shaded area represents its 95% confidence interval. The other three lines are estimated from observations at the DMA-year-month level. All regressions additionally include the mean temperature and precipitation in given and previous months. The underlying coefficients are presented in Table S9.

### Projected impacts of solar geoengineering

We project the first estimates of how reduced incoming sunlight by solar radiation management will affect the suicide rate. We first disentangle the effects of an anticipated shift in sunlight (climate effect) and an unanticipated shock to sunlight exposure (weather effect) (Methods). Since the future impacts depend solely on the anticipated climate effect, we project the future impacts of sunlight shift by solar geoengineering using the parameters for the climate effect (35).

To arrive at an “excess” incidence of suicides due to reduced sunlight, we compute the negative radiative forcing gap, or the amount of reduction in solar irradiance necessary to keep the global temperature rise below 1.5 °C, under business-as-usual and various emission reduction scenarios of CO_2_ emissions. We then multiply this change in solar insolation by our estimated effect of sunlight on the suicide rate and projected population growth under the assumption that the impacts of an anticipated shift in sunlight in the past decades can be extrapolated to the period of 2030–2100.

Overall, our estimates suggest that solar insolation needs to be reduced by up to 1.69% in 2100 under the business-as-usual scenario, which will cause a cumulative 2.23 (95% CI: 1.26, 3.18) thousand additional suicides by 2100 (Figure 5, Table S12). In contrast, temperature is projected to rise by 4 °C in 2100 under the business-as-usual scenario. By reducing the temperature rise by about 2.5 °C to achieve the goal of a 1.5 °C temperature rise, we project that 5.75 (95% CI: 1.23 10.69) thousand suicides will be averted. Thus, the effects of insufficient sunlight will offset about 38.8% (95% CI: 27.8, 163.5) of the suicides averted by temperature fall. The resulting net reduction in the cumulative suicides by 2100 will be 3.52 (95% CI: −0.781, 7.72) thousand (Table S12). Thus, solar geoengineering can indeed increase the incidence of suicides when the effects of insufficient sunlight more than offset the effects of temperature (Figure S5).

**Figure 5:**
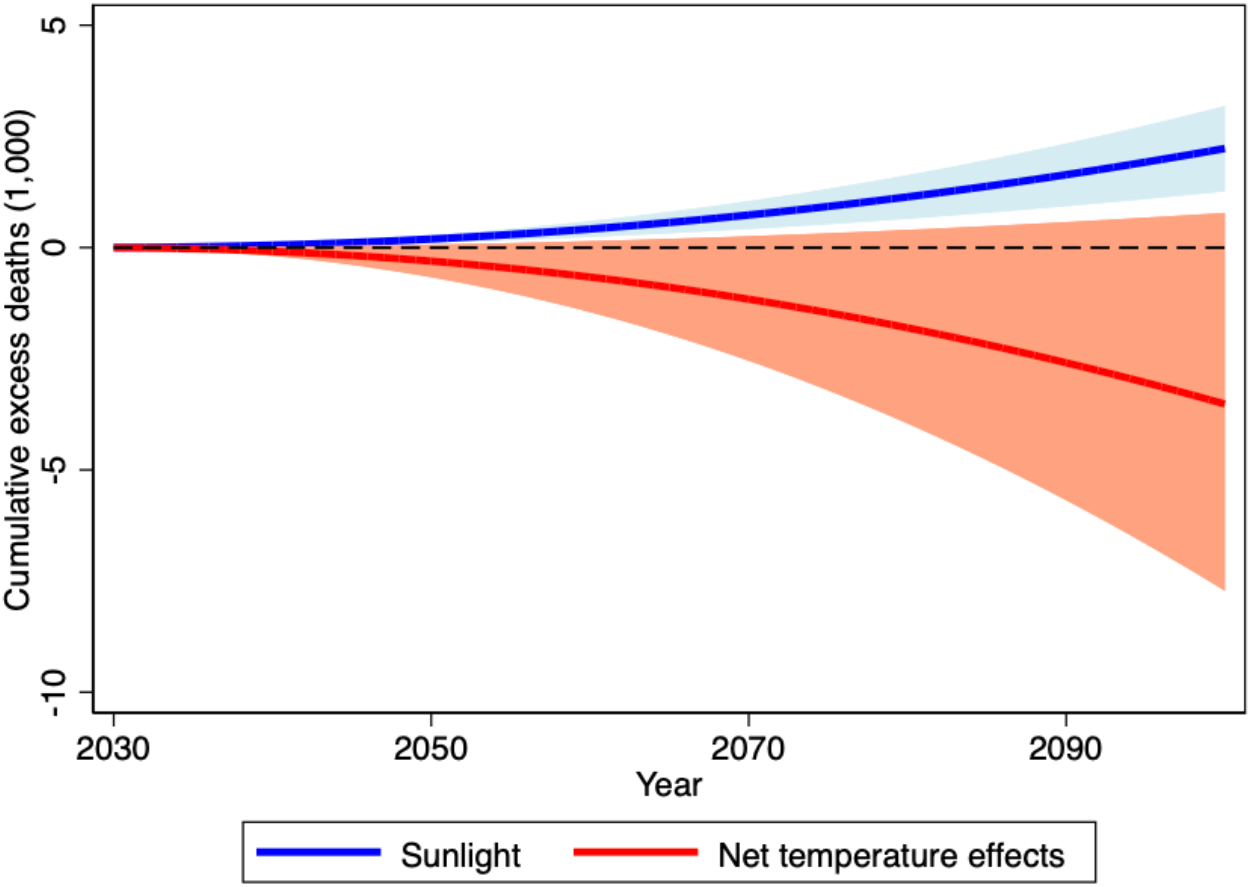
Projected impacts of solar radiation management on suicides, 2030-2100. *Notes*: This figure plots the projected impacts of solar radiation management on suicides to keep the global temperature rise below 1.5 °C between 2030 and 2100. The blue line indicates the cumulative excess suicide deaths due to the negative radiative forcing, and the shaded area represents the 95% CI. The red line indicates the net increase in suicide deaths after accounting for averted suicides due to temperature fall, and the shaded area represents the 95% CI.

## Discussion

This study assessed whether and to what extent sunlight exposure affects the suicide rate and the implications for solar geoengineering. Compared to previous research, our empirical approach has three novel advantages. First, we employed large-scale data including approximately 445,000 observations at the county-by-month level over nearly three decades. Second, we used solar insolation per county-by-month as a direct measure of exposure to sunlight, which is conceptually more valid and precise than commonly used measures such as the duration of daylight. Third, our estimation approach minimizes confoundedness by controlling for geographic and seasonal characteristics specific to a particular county in a particular month of the year. Given that the amount of sunlight is plausibly random after adjusting for factors related to sunlight itself, e.g. temperature and precipitation, we identified the causal effect of insufficient sunlight on increasing suicides. In contrast to our findings, a handful of studies using solar insolation or irradiation as the exposure measure relied exclusively on a time-series analysis and showed a positive association with suicide (30). Using our data, we found that such a positive association arises because time-series data suffer from a severe bias due to the seasonality effect (Table S8). In contrast, we provided counter evidence because our analysis, based on longitudinal data, adequately adjusts for geographic and seasonal variations related to solar insolation.

Our main findings suggest that suicide rates increase by 6.99% (95% CI: 3.86, 10.13) as sunlight in given and previous months decreases by one standard deviation. This amount of change in sunlight is approximately equivalent to the difference in the average state-level sunlight between Vermont at the lowest level of sunlight and Arizona at the highest level of sunlight during our study period of 1979–2004. We found few heterogeneities in the sunlight-suicide relationship by county characteristics or over time. Our additional analysis indicates that sunlight is negatively related to the volume of internet searches for depressive language on Google, indicating that sunlight affects suicide by affecting individuals’ mental well-being.

Let us compare our estimated impact of sunlight on the suicide rate with the impacts of other interventions in previous studies. Figure 6 describes the amount of changes in sunlight in both percentage and standard deviation that brings about the equivalent impacts by other factors such as air pollution (25), temperature (24), firearm regulations (36), national suicide prevention programs (37), celebrity suicides (38), a higher unemployment rate (39), and COVID-19 (40). For example, a 0.34 standard deviation, or 13.3%, decrease in sunlight brings about the equivalent effect of a 10-*μg*/*m*^3^ increase in PM_10_ on suicide. Overall, we find that a 1 to 2 standard deviation change in sunlight generates the equivalent impact of other interventions, revealing sunlight as a major risk factor in the incidence of suicide.

**Figure 6:**
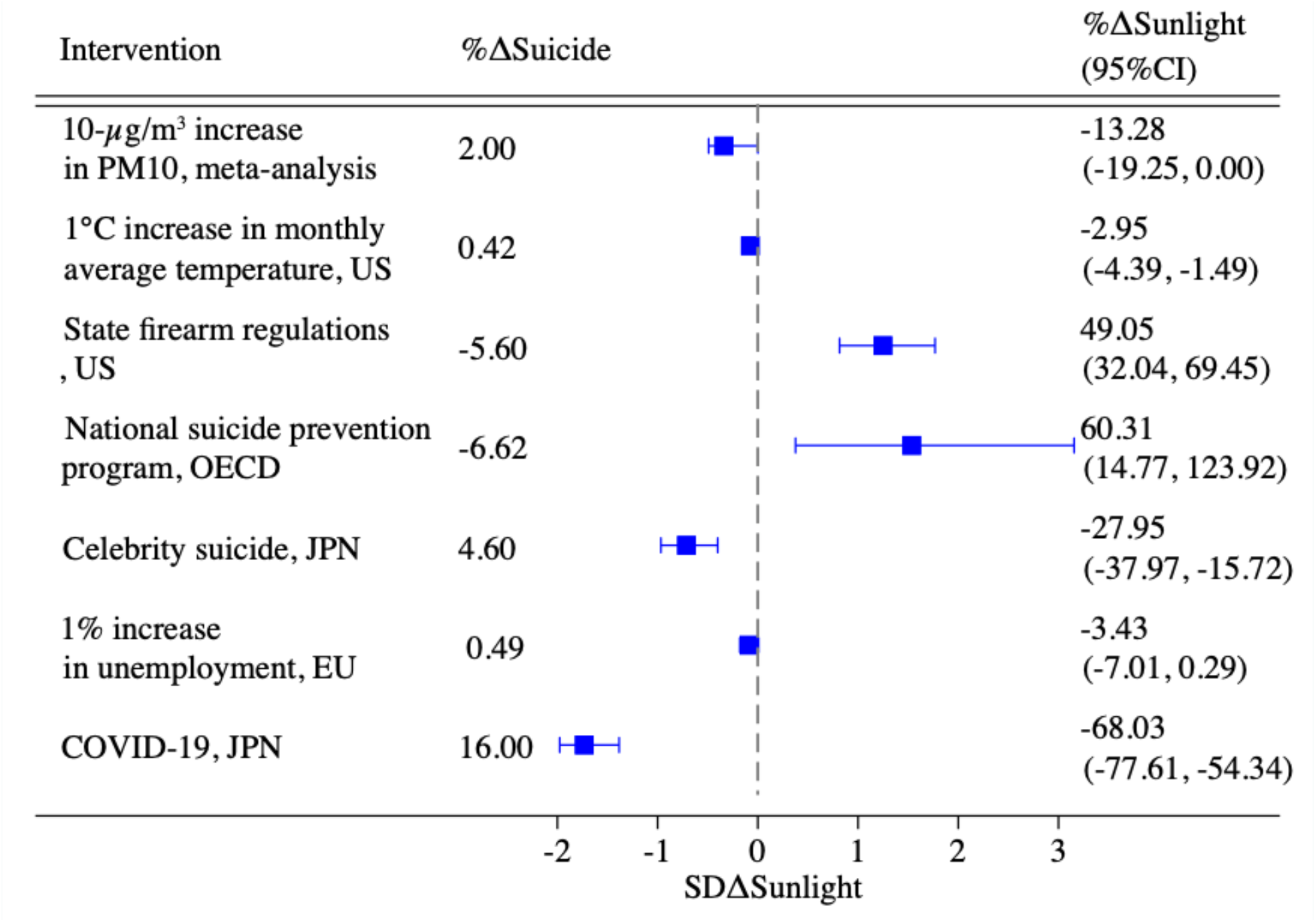
Changes in sunlight required to achieve the equivalent impacts of other interventions. *Notes*: This figure describes the amount of changes in sunlight in both percentage and standard deviation (SD) that is required to achieve the equivalent impacts of other interventions estimated in previous studies (25,24,36–40). “Intervention” describes the type of intervention, “%ΔSuicide” describes the percentage change in the suicide rate by the intervention, each dot describes the change in the standard deviation of sunlight that is required to achieve the same magnitude of the impact as the intervention, along with its associated 95% CI, and “%ΔSunlight” indicates the percentage change in sunlight that is required to achieve the same magnitude of impact as the intervention. The underlying parameters are reported in Table S11.

Our projection suggests that reducing solar radiation to keep the global temperature rise below 1.5 °C, as currently targeted by the Paris Agreement, could result in 1.26–3.18 thousand additional suicides (95% CI) by 2100, and the net of the temperature reduction effects is projected to cause 0.781 to −7.72 additional suicides by 2100. Thus, solar geoengineering can indeed increase the incidence of suicides when the effects of insufficient sunlight more than offset the effects of temperature.

Our study involves two limitations. First, our data are obtained only from the (contiguous) U.S. states. This limits the geographical variation in the amount of solar insolation and characteristics of populations, and the observed relationships may differ if data include additional observations with solar insolation beyond the range observed in our sample or in other countries. Thus, the findings should be carefully interpreted for generalizability, especially for places at extremely high or low latitudes. From a distributive justice perspective, it would also be useful to explore potential differences in the effects of solar geoengineering between developed and developing countries.

Second, more importantly, our findings offer limited insight into the mechanism(s) whereby greater exposure to sunlight reduces suicide risk. The relationship between sunlight and online depressive language is suggestive, yet it is still unclear why and how reducing exposure to sunlight worsens people’s mental conditions. The mechanisms may well be biological and/or behavioral. Biologically, insufficient sunlight is known to cause disrupted sleep, impeded neurotransmissions of serotonin, vitamin D deficiency, and the overproduction of melatonin levels (16–18), all or some of which may adversely affect mental health and increase suicide risk. The behavioral component may stem from the association of greater sunlight with more physical activity and more social interactions, both of which can reduce feelings of sadness and isolation. Such a behavioral linkage is likely for individuals in places with low levels of sunlight who would spend more time outdoors on sunnier days (holding temperature constant). On the other hand, individuals in places with high levels of sunlight would spend more time indoors when sunlight is greater (41), which would lead to a positive association, or at least a weaker negative association, between sunlight and suicides. Thus, such a behavioral mechanism contradicts our finding that the effect is strongest in the top decile areas. In addition, the substantial cumulative impacts from the previous month suggest the mechanism leading to a suicide attempt is a dynamic process over months. In contrast, temperatures and air pollution have immediate effects on suicides, as aggressive emotions or inflammation of the nervous system trigger self-harm attempts (42). Future research is warranted to discover the precise underlying mechanism(s) at work.

This study has important policy implications. Most research on sunlight has focused on adverse health consequences such as skin cancer, resulting in the widespread awareness and current public health advice to reduce sunlight exposure. However, increased exposure to sunlight has a preventive impact on suicide along with other health benefits. Our findings offer key insights that supplemental policy remedies, such as suicide prevention programs or mental health assistance programs, may mitigate some of the adverse impacts of solar geoengineering. It is thus vital for climate policy remedies, such as geoengineering, to evaluate and attempt to balance the potential benefits and harms of solar radiation.

## Data Availability

All data produced in the present study are available upon reasonable request to the authors.

## Materials and Methods

### Data

Our data on suicide comes from Burke et al. (24), which reports the age-adjusted suicide rates at the county-month level based on the Multiple Cause of Death Mortality Data from the National Vital Statistics System between 1968 and 2004. The data contained all counties until 1988 and counties with more than 100,000 residents after 1989. The data period necessarily ended in 2004 because county identifiers were not reported in the public-use data after 2004. The data also included the monthly average temperature and total precipitation from PRISM. See more details on how these variables were constructed in Burke et al. (24).

We combined the suicide data with average daily solar insolation data at the county-by-month level in kilojoules per square meter (KJ/m^2^) from the North America Land Data Assimilation System Daily Sunlight data compiled by the U.S. Centers for Disease Control and Prevention (43). Solar insolation is the amount of incident solar radiation energy received from the Sun per unit surface of the Earth over a specified period of time. Many factors determine how much sunlight reaches the surface, such as the solar zenith angle, the variable distance from the earth from the sun, day length, weather conditions, atmospheric aerosols levels, and the solar activity. The original data covered the 48 contiguous states plus the District of Columbia from 1979 to 2011. Thus, the resulting data after merging sunlight and suicide information spanned the period from 1979 to 2004 across (unbalanced) 3,107 counties in the 48 contiguous states and the District of Columbia.

To explore the potential heterogeneities in the effects of sunlight on suicide rates by various county characteristics, we drew annual county income data from the U.S. Bureau of Economic Analysis, deflated by the GDP deflator, data on the state-level average adoption of air conditioning between 1979 and 2004 from Barreca et al. (34), and state-level gun ownership data from Okoro et al. (36). The suicide rates by gender and methods of suicide are from Burke et al. (24).

We analyzed the impacts of sunlight exposure on the volume of searches made on Google using Google Trends. Google Trends reports the search data at several geographical levels, and two sets of the regions useful in our context were the state and Designated Marketing Area (DMA), whereas the county data were unavailable. A DMA is a geographically delineated media market, in which people receive the same television and radio options. There are 210 DMAs in total across the U.S. Google Trends reports normalized search interests, rather than raw search volumes, on a scale of 0 to 100, by comparing the search volumes in the respective time and region relative to the highest point under the selected condition. For example, an index of 50 at a given time (e.g., month, week, or day) represents half the volume of searches with the index value of 100 at the same time.

Given the restrictions on the number of regions and keywords that could be included for each search, and because the data dated back to 2004, each round of our data collection involved the words “depression” and one other keyword from the list of depressive language below for 2004–2011 by each region. For example, we selected “depression” and “suicide” as search terms, 2004–2011 as the time range, and California as the location. We selected the word “depression” as the reference word because this yielded greater search volumes than most other keywords, and in this way the Google Trends values for each keyword were comparable within each region. The set of depressive language followed Burke et al. (24) and included: addictive, alone, anxiety, appetite, attacks, bleak, depress, depressed, depression, drowsiness, episodes, fatigue, frightened, lonely, nausea, nervousness, severe, sleep, suicidal, suicide, and trapped. We summed up Google Trends values for all these keywords at the region-by-year-by-month level and again normalized them to the highest value within the region with a scale of 0 to 100 to construct the overall normalized search interests in depressive language. Note that while the constructed Google Trends values were not comparable across regions, e.g., 100 in region A is not comparable to 100 in region B, the inclusion of region fixed effects addressed this issue.

### The effects of sunlight exposure on suicide rate

The main analysis estimated the following distributed lag models that included the lags and leads of each environmental factor, using ordinary least squares:

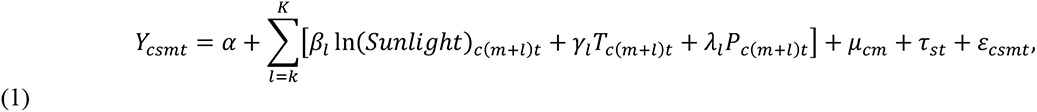

where the main outcome variable is the suicide rate per 100,000 population in county *c* in state *s* in month *m* of year *t*. The main independent variable of interest, ln(*Sunlight*), is the log of average daily solar insolation (in KJ/m^2^), *T* denotes the monthly average temperature, and *P* denotes the monthly precipitation. The county-by-month fixed effects, *μ*_*cm*_, controlled for unobserved seasonality effects at the county-month level, such as the daylight duration or the school calendar, whereas the state-by-year fixed effects, *τ*_*st*_, accounted for any time-varying factors common across counties within a given state, such as economic conditions, agricultural production, poverty rates, and gun ownership. Thus, the parameters of interest, *β*_*l*_’s, were estimated based on the random variation in the amount of sunlight between, for example, August 2000 in a particular county and August 2001 in the same county. Following the convention, the regressions were weighted by county population, as the suicide rate is more precisely estimated with a larger population. Further, we clustered the standard errors at the county level to correct heteroskedasticity in the error term, *ε*_*csmt*_, and allowed for correlations between observations within clusters. We also tested alternative levels of clustering, such as the state level and two-way clustering at the county and year level, to account for spatial correlation in sunlight and showed that the results are virtually the same (Table S4).

The identification assumption is that such within-county-month variation in the amount of sunlight was uncorrelated with any other factors that affected the suicide rate. Such assumption is plausible because the amount of sunlight was determined by random fluctuations in climatic and meteorological conditions, after adjusting for factors related to sunlight itself, such as temperature, precipitation, the average sunlight in a given county-by-month, and year-specific shocks common within a given state. Air pollution may be another environmental factor that is related to both sunlight and suicides. Unfortunately, reliable data on air pollution is available only for recent years, e.g., after 2000 (44). As a robustness check, we confirmed that the estimated effect of sunlight is unchanged with and without controlling for the PM2.5 concentrations in ambient air in 2000–2004, the period during which we have both air pollution and suicide data (Table S5).

The distributed lag models allowed us to examine whether insufficient sunlight exposure caused excess suicides or simply hastened suicides that would have occurred later anyway, the so-called harvesting effect. Each parameter, *ω*_*l*_ ∈ (*β*_*l*_, *γ*_*l*_, *λ*_*l*_) for *l* ∈ [*k, K*], can be interpreted as the effects of sunlight, temperature, and precipitation, respectively, in each month with lags and leads of *l* month. For example, *ω*_0_ indicates the effect of a given month’s environmental factor, *ω*_−1_ the previous month’s factor, and *ω*_1_, the following month’s factor. A finding of *β*_*l*_ < 0 for *l* < 0 indicates the lagged impacts of sunlight exposure in the previous *l* month on the current suicide rate, whereas a finding of *β*_*l*_ > 0 for *l* < 0 indicates a displacement effect, where insufficient sunlight in a particular month hastened suicides that would have occurred anyway in a −*l* month later. Thus, the overall effect of sunlight in a given month was given by 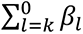. We expected *ω*_*l*_ =0 for *l* > 0 as a placebo test since a future amount of sunlight should not have a causal impact on the incidence of suicide in a given month.

Since the main model is the level-log model, the interpretation of the coefficient is that a 1% increase in sunlight increases the suicide rate by *β*_*l*_/100. To allow for potential nonlinear effects of sunlight, we also considered a 3^rd^ order polynomial function of average daily sunlight, where the main independent variable was replaced with 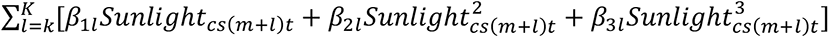. The temperature and precipitation were included as a linear function based on evidence presented by Burke et al. (24). As a further robustness check, we also considered a nonparametric binned model, where the main independent variable was replaced with 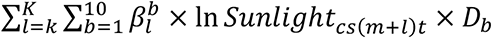, where *D*_*b*_ was an indicator variable for each decile bin *b*. Based on the results from the main analysis and to ensure greater statistical power, we considered *l* ∈ [−1,0], whereas extending the period did not alter the conclusion.

A concern may arise about the endogeneity of temperature to sunlight, making it a “bad control” (45). However, our main model includes temperature as a control for two reasons. One is that our goal is to estimate the effects of sunlight itself free from the effects of temperature. Because increased sunlight raises temperature, a model that did not control for temperature would estimate the overall impacts of sunlight on suicides resulting from two offsetting channels; reductions in suicides due to increased sunlight itself and increases in suicides due to higher temperatures. Thus, the inclusion of temperature as a control helps us isolate the impacts of sunlight on suicides net of temperature effects. Indeed, a model that does not control for temperature understates the effects of sunlight itself on suicides (Table S6). Second, the effect of sunlight on temperature is small, rendering the problem of endogeneity negligible. For example, the county-by-month and state-by-year fixed effects explain 97.27% of the overall variation in temperature, whereas precipitation only accounts for an additional 0.01 percentage point explanatory power, and sunlight adds 0.03 percentage points (Table S7). We also showed that a 1 standard deviation decrease in sunlight leads to a 0.144 standard deviation decrease in temperature. Thus, the overall variation in temperature that is explained by sunlight is small.

### Heterogeneities in the effects of sunlight

To estimate the potential heterogeneous effects of sunlight exposure on the suicide rate by various county- or state-level characteristics, i.e., sunlight, temperature, income, air conditioning ownership, and gun ownership, we first computed the population-weighted county- or state-level median characteristics and ran a regression for each characteristic:

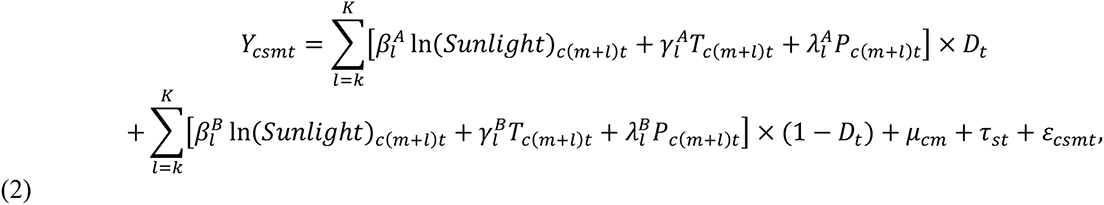

where *D* is an indicator variable for being above the median characteristics. Then, 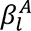 indicates the effects of sunlight in counties with above-median characteristics and 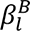 indicates the effects for counties with below-median characteristics.

### Adaptation over time

We explored how individuals adapt to variations in sunlight over time. We first applied a model comparable to the main analysis by interacting each environmental factor with a dummy variable for the respective year.

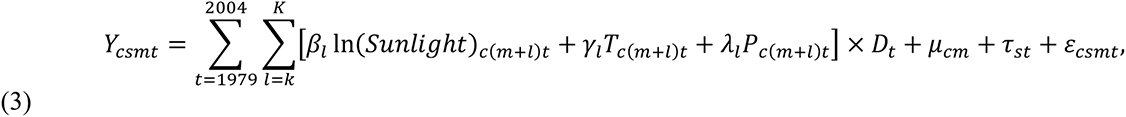

where *D*_*t*_ is an indicator variable for each year *t*. Based on the results from the main analysis and to ensure greater statistical power, we consider *l* ∈ [−1,0], whereas extending the period does not alter the conclusion.

### Sunlight and depressive language in Google Trends

We estimated the effects of monthly average sunlight on Google searches containing depressive language by regressing the following fixed-effects model using the ordinary least squares:

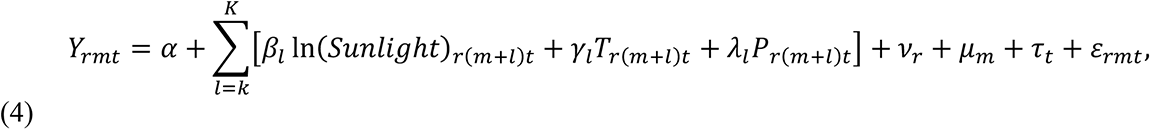

where *Y*_*rmt*_ is the Google Trends index in region *r* ∈(state, DMA), month *m*, and year *t*. Based on the results from the main analysis and to ensure greater statistical power, we considered *l* ∈ [−1,0], whereas extending the period did not alter the conclusion. The regressions were weighted by regional population, and the standard errors were clustered at the regional level. Since the time framework for Google Trends data did not overlap with that of the main analysis, we additionally obtained population data from the U.S. Census. We also obtained the temperature and precipitation information from NOAA’s Global Historical Climatology Network - Daily.

As robustness checks, we included a set of alternative fixed effects. For example, with the DMA-level data, we included state-by-year and state-by-month fixed effects or state-by-month and state-specific trends, as in the main analysis. Note that many DMAs cross multiple states. In such a case, we selected the state that was referenced in the DMA name as the primary state of affiliation.

As further robustness checks, we estimated the same models for a different subset of depressive language terms and found similar results (Table S9).

### Projected impacts of solar geoengineering

The estimated effects of sunlight on suicides thus far include both the effects of an anticipated shift in sunlight over time, i.e., climate effects, and the effects of an unanticipated shock to sunlight exposure, i.e., weather shocks. For example, individuals in areas with greater sunlight may shelter themselves from the increased harms of sunlight exposure due to, for instance, ozone depletion, by shifting their work from outdoors to indoors over time, whereas current daily activities, such as work and school, are less responsive to day-to-day fluctuations in sunlight. In this case, the marginal effects of an unanticipated shock are likely to outweigh the marginal effects of an anticipated shift over time (35), with the result that the projected impacts of solar geoengineering based on weather shocks overstate the actual impacts of the climate effect. Thus, we adopted a model that explicitly disentangled these two effects as follows:

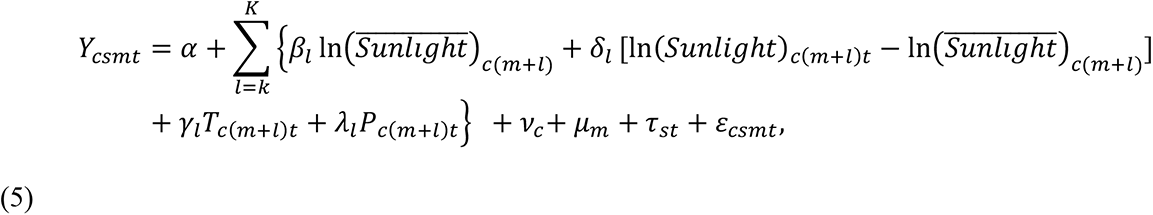

where we separately controlled for the county-month average sunlight in our study period, 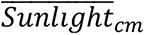, and the difference between the observed sunlight and long-term average sunlight for each county and month. Since the county-by-month fixed effects would be multicollinear with the county-month’s average sunlight, we separately controlled for county fixed effects, *v*_*c*_, and month fixed effects, *μ*_*m*_. The county fixed effects controlled for permanent differences in county characteristics that affect the suicide rate, whereas the month fixed effects controlled for seasonality patterns across months in the suicide rate. Additionally, we controlled for state-by-year fixed effects to control for transitory shocks to the suicide rate that are common within a particular year of the state.

The effect of an anticipated sunlight shift over time is captured by the 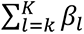 coefficients, which are identified from within-county variation in the average sunlight in each month, after controlling for national seasonality. For example, sunlight may be stronger in August than in July in County A much more so than in County B, leading people in County A to spend more time indoors than those in County B. In contrast, the effects of an unanticipated sunlight shock in a given month of a particular year is captured by the 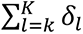 coefficients, which are identified from a transitory deviation from the anticipated level of sunlight in a specific month.

Then, using the parameter based on the climate effect, we projected how the reduction of sunlight due to solar radiation management will affect the incidence of suicide between 2030 and 2100. We assumed that the cumulative CO_2_ emissions in 2030 would be 700 Gt (46) and would grow at the rate of 47 Gt/year under a business-as-usual scenario thereafter (47). Temperature is already set to be above 1 °C from preindustrial times in 2015 (48) and is projected to increase by 1 °C for every 1,300 Gt CO_2_ emissions (47). Thus, the cumulative CO_2_ emissions must stay at 650 Gt from 2015 onward to keep the temperature rise below 1.5°C or 1,300 Gt for the temperature to be below 2°C. The equivalent radiative forcing amount is taken from the equilibrium climate sensitivity of approximately 0.8 °C/(W/m^2^) (49). Together, these parameters imply the amount of radiative forcing that is required to offset the temperature rise due to 1 Gt of CO_2_ is 9.6 × 10^−4^ (W/m^2^)/Gt(CO_2_) (47).

Using these parameters, the cumulative number of excess suicides due to the reduction in sunlight required to meet the goal of keeping global temperature rise below 1.5 °C between 2030 and 2100 can be obtained by:

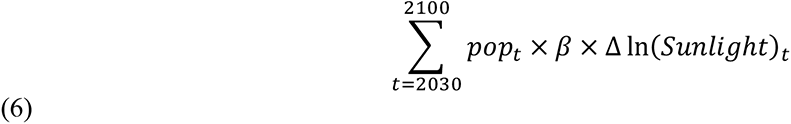

where *pop*_*t*_ is the projected U.S. population in year *t* in hundred thousand from United Nations (50), *β* is the estimated climate impact of sunlight on the suicide rate (*β* =−0.078, 95%CI: −0.112, −0.044) from Table S10 Column (1)), and Δln (*Sunlight*) is the log point change of the implied negative radiative forcing gap to achieve the temperature limit in each year from the mean (set to be zero if the radiative forcing gap is positive since radiative forcing need not be increased to meet the temperature limit). For example, in 2030, the cumulative CO_2_ emissions will already exceed the remaining CO_2_ budgets of 650 Gt by 50 Gt to keep the temperature rise below 1.5 °C. The implied negative radiative forcing gap is then 0.048 W/m^2^ (=50 × 9.6 × 10^−4^). This is equivalent to a 4.4172 KJ/m^2^ reduction in the daily solar insolation, which is a 0.0002526 log point reduction from its mean, i.e., an approximately 0.025% reduction in daily solar insolation. Finally, to arrive at the annual increase in suicides, we multiplied the number by the projected population and by 12 months.

We conducted a similar analysis to compute “averted” suicides due to reduced temperature to meet the goal of keeping global temperature rise below 1.5 °C between 2030 and 2100 by:

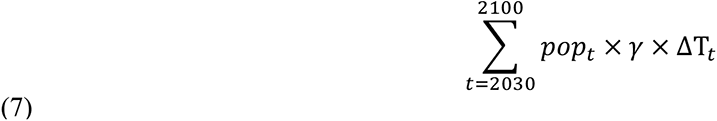

where ΔT_*t*_ is the change in temperature from the business-as-usual scenario to achieve the temperature limit (again set to be zero if the temperature is below the limit). For example, in 2030, the temperature will exceed the 1.5°C limit by 0.038°C. We then multiply 0.038 by the estimated impacts of temperature in given and previous month on the suicide rate (*γ* =0.00134 (95% CI: 0.000287, 0.00248) from Gammans (35)) to obtain the averted suicide rates. We take the temperature effects from Gammans (35) because our unbalanced data do not allow us to compute the county-month average temperature in our study period. Nonetheless, the estimated effects using the county-month average temperature within our data produce very similar impacts. To arrive at the annual reductions in suicides, we multiplied the number by the projected population and by 12 months.

To compute the net effects of sunlight and temperature on the incidence of suicides, we randomly drew *β* and *γ* from its estimated distribution and projected their cumulative impacts in 2100. We repeated the process by 10,000 times to arrive at the 95% CI.

## Supplementary Materials

**Table S1:** Summary Statistics

| variable | N | Mean | Std |
| --- | --- | --- | --- |
| Suicide rate (per 100K pop) | 444,861 | 0.955 | 0.967 |
| Sunlight (KJ/m<sup>2</sup>/day) | 444,861 | 16,422.91 | 6,449.22 |
| Population | 444,861 | 143,376 | 384,245 |
| Temperature (C) | 444,861 | 13.822 | 9.120 |
| Precipitation (mm) | 444,861 | 0.073 | 0.059 |
| Suicide rate, male nonviolent | 444,861 | 0.210 | 0.578 |
| Suicide rate, male violent | 444,861 | 1.171 | 1.581 |
| Suicide rate, female nonviolent | 444,861 | 0.159 | 0.458 |
| Suicide rate, female violent | 444,861 | 0.215 | 0.633 |
| Real income (USD) | 439,202 | 31347.5 | 10047.4 |
| Gun ownership | 444,861 | 0.309 | 0.120 |

*Notes:* This table reports the summary statistics of the main variables. Each column reports the variable name, the number of observations, the mean value weighted by population (except for population, where the mean is the arithmetic average), and the standard deviation, respectively. The sample is restricted to what is used in the main analysis. The level of observations is at the county-by-month level.

**Table S2:** The Effects of Sunlight on Suicide Rate

|  | (1) | (2) | (3) | (4) | (5) | (6) |
| --- | --- | --- | --- | --- | --- | --- |
| $\ln(\text{Sunlight})_0$ | -0.049**<br>(0.022) | -0.052**<br>(0.022) | -0.045*<br>(0.025) | -0.060***<br>(0.022) | -0.043*<br>(0.023) | |
| $\ln(\text{Sunlight})_{-1}$ | -0.085***<br>(0.024) | -0.083***<br>(0.024) | -0.086***<br>(0.025) | -0.093***<br>(0.024) | -0.077***<br>(0.024) | |
| $\ln(\text{Sunlight})_{-2}$ | -0.003<br>(0.022) | 0.001<br>(0.021) | -0.002<br>(0.023) | -0.008<br>(0.021) | 0.003<br>(0.022) | |
| $\ln(\text{Sunlight})_1$ | 0.010<br>(0.024) | 0.006<br>(0.023) | 0.002<br>(0.025) | -0.004<br>(0.023) | 0.017<br>(0.024) | |
| $\text{Temp}_0$ | 0.008***<br>(0.001) | 0.009***<br>(0.001) | 0.009***<br>(0.001) | 0.008***<br>(0.001) | 0.008***<br>(0.001) | 0.008***<br>(0.001) |
| $\text{Temp}_{-1}$ | -0.002**<br>(0.001) | -0.001<br>(0.001) | -0.001<br>(0.001) | -0.002**<br>(0.001) | -0.002**<br>(0.001) | -0.002**<br>(0.001) |
| $\text{Temp}_{-2}$ | 0.000<br>(0.001) | 0.001<br>(0.001) | 0.001<br>(0.001) | 0.000<br>(0.001) | -0.000<br>(0.001) | 0.000<br>(0.001) |
| $\text{Temp}_1$ | 0.001<br>(0.001) | 0.002*<br>(0.001) | 0.002<br>(0.002) | 0.001<br>(0.001) | 0.001<br>(0.001) | 0.001<br>(0.001) |
| $\text{Precip}_0$ | -0.043<br>(0.032) | -0.012<br>(0.034) | -0.001<br>(0.036) | -0.032<br>(0.032) | -0.024<br>(0.033) | -0.043<br>(0.033) |
| $\text{Precip}_{-1}$ | -0.014<br>(0.036) | 0.032<br>(0.038) | 0.044<br>(0.041) | 0.011<br>(0.037) | 0.003<br>(0.036) | -0.013<br>(0.036) |
| $\text{Precip}_{-2}$ | -0.013<br>(0.034) | 0.042<br>(0.037) | 0.046<br>(0.040) | 0.016<br>(0.032) | 0.003<br>(0.034) | -0.011<br>(0.034) |
| $\text{Precip}_1$ | 0.019<br>(0.032) | 0.044<br>(0.031) | 0.065*<br>(0.033) | 0.029<br>(0.032) | 0.040<br>(0.033) | 0.020<br>(0.032) |
| $\text{Sunlight}_0$ | | | | | | -0.020<br>(0.013) |
| $\text{Sunlight}_0^2$ | | | | | | 0.001<br>(0.001) |
| $\text{Sunlight}_0^3$ | | | | | | -0.000<br>(0.000) |
| $\text{Sunlight}_{-1}$ | | | | | | -0.011<br>(0.013) |
| $\text{Sunlight}_{-1}^2$ | | | | | | -0.000<br>(0.001) |
| $\text{Sunlight}_{-1}^3$ | | | | | | 0.000<br>(0.000) |
| $\text{Sunlight}_{-2}$ | | | | | | 0.002<br>(0.013) |
| $\text{Sunlight}_{-2}^2$ | | | | | | 0.000<br>(0.001) |
| $\text{Sunlight}_{-2}^3$ | | | | | | -0.000<br>(0.000) |
| $\text{Sunlight}_1$ | | | | | | 0.010<br>(0.013) |
| $\text{Sunlight}_1^2$ | | | | | | -0.001<br>(0.001) |
| $\text{Sunlight}_1^3$ | | | | | | 0.000<br>(0.000) |
| Constant | 2.088***<br>(0.461) | 2.025***<br>(0.408) | 2.032***<br>(0.454) | 17.855<br>(36.095) | 1.828***<br>(0.477) | 1.080***<br>(0.158) |
| adj. $R^2$ | 0.11 | 0.11 | 0.11 | 0.11 | 0.16 | 0.11 |
| $\beta_0 + \beta_{-1}$ | -0.134 | -0.136 | -0.131 | -0.152 | -0.120 | -0.141 |
| $se(\beta_0 + \beta_{-1})$ | 0.031 | 0.030 | 0.034 | 0.030 | 0.031 | 0.047 |
| $p(\beta_0 + \beta_{-1})$ | 0.000 | 0.000 | 0.000 | 0.000 | 0.000 | 0.003 |
| Fixed effects | county × month<br>+ state × year | county × month<br>+ year | county × month<br>+ year × month | county × month<br>+ year + state trend | county × month<br>+ county × year | cubic<br>polynomial |

*Notes:* The dependent variable is the suicide rate per 100,000 population. The mean of the dependent variable is 0.955, and the mean of sunlight is 16422.91 KJ/m<sup>2</sup>. The level of observations is at the county-month. The sample size is 444,861 across 3,107 counties. At the bottom of the table, we report the linear combination of $\beta_0 + \beta_{-1}$ , which represents the cumulative effects of sunlight in given and previous months as well as the associated standard errors and the $p$ -values. The corresponding value in Column (5) has the same interpretation, which is the changes in the suicide rate by a 100% increase in sunlight from its mean value. All regressions are weighted by county population. The standard errors clustered at the county level are reported in the parentheses. \*\*\* $p < 0.01$ , \*\* $p < 0.05$ , \* $p < 0.1$

**Table S3:** Robustness to Alternative Outcomes

|  | (1) | (2) | (3) | (4) |
| --- | --- | --- | --- | --- |
|  | ln(rate+1) | ln(count+1) | IHS | Count |
| ln(Sunlight)<sub>0</sub> | -0.049**<br>(0.022) | -0.015<br>(0.011) | -0.038**<br>(0.015) | -0.052**<br>(0.024) |
| ln(Sunlight)<sub>-1</sub> | -0.085***<br>(0.024) | -0.019*<br>(0.011) | -0.055***<br>(0.016) | -0.089***<br>(0.025) |
| ln(Sunlight)<sub>-2</sub> | -0.003<br>(0.022) | -0.015<br>(0.011) | -0.005<br>(0.015) | -0.007<br>(0.023) |
| ln(Sunlight)<sub>1</sub> | 0.010<br>(0.024) | 0.020*<br>(0.011) | 0.004<br>(0.016) | 0.009<br>(0.026) |
| Temp<sub>0</sub> | 0.008***<br>(0.001) | 0.003***<br>(0.000) | 0.005***<br>(0.001) | 0.008***<br>(0.001) |
| Temp<sub>-1</sub> | -0.002**<br>(0.001) | -0.000<br>(0.000) | -0.001**<br>(0.001) | -0.002**<br>(0.001) |
| Temp<sub>-2</sub> | 0.000<br>(0.001) | 0.000<br>(0.000) | 0.000<br>(0.001) | 0.000<br>(0.001) |
| Temp<sub>1</sub> | 0.001<br>(0.001) | 0.000<br>(0.000) | 0.001<br>(0.001) | 0.001<br>(0.001) |
| Precip<sub>0</sub> | -0.043<br>(0.032) | -0.004<br>(0.015) | -0.028<br>(0.021) | -0.041<br>(0.034) |
| Precip<sub>-1</sub> | -0.014<br>(0.036) | 0.001<br>(0.015) | -0.006<br>(0.025) | -0.013<br>(0.038) |
| Precip<sub>-2</sub> | -0.013<br>(0.034) | -0.008<br>(0.016) | -0.009<br>(0.022) | -0.014<br>(0.035) |
| Precip<sub>1</sub> | 0.019<br>(0.032) | 0.014<br>(0.015) | 0.017<br>(0.022) | 0.024<br>(0.033) |
| ln(population) |  | 0.430***<br>(0.034) |  |  |
| Constant | 2.088***<br>(0.461) | -3.828***<br>(0.387) | 1.595***<br>(0.309) | -10.232***<br>(0.484) |
| adj. R<sup>2</sup> | 0.11 | 0.73 | 0.19 |  |
| $\beta_0 + \beta_{-1}$ | -0.134 | -0.034 | -0.093 | -0.141 |
| se($\beta_0 + \beta_{-1}$) | 0.031 | 0.015 | 0.020 | 0.032 |
| $p(\beta_0 + \beta_{-1})$ | 0.000 | 0.022 | 0.000 | 0.000 |

*Notes:* This table reports the estimates from various alternative specifications. In particular, Column (1) uses the log of the suicide rate per 100,000 population plus one as the dependent variable, Column (2) uses the log of the suicide count plus one as the dependent variable, Column (3) uses the inverse hyperbolic sine (IHS) of the suicide rate as the dependent variable, and Column (4) applies the Poisson model to the suicide count as the dependent variable. The sample size is 444,861 in Columns (1)–(3) and 364,778 in Column (4), as Poisson pseudo-maximum likelihood regressions with multi-way fixed effects additionally detects and drops separated observations. All regressions control for county-month and state-year fixed effects. Regressions are weighted by population in Columns (1) and (3), while Column (2) additionally controls for the log of population, and Column (4) is estimated by the Poisson regression that includes the population as an exposure variable. The interpretations of the bottom parameters are the same as Table S2.
\*\*\* $p < 0.01$ , \*\* $p < 0.05$ , \* $p < 0.1$

**Table S4:** Robustness to alternative levels of clustering the standard errors

|  | (1) | (2) | (3) | (4) |
| --- | --- | --- | --- | --- |
| $\beta_0 + \beta_{-1}$ | -0.134*** | -0.134*** | -0.134*** | -0.134*** |
| $se(\beta_0 + \beta_{-1})$ | (0.031) | (0.031) | (0.041) | (0.028) |
| $p(\beta_0 + \beta_{-1})$ | [0.000] | [0.000] | [0.003] | [0.000] |
| Clustering | county | county<br>+ state-year | county<br>+ year | state |

*Notes:* This table tests the robustness of the effects of sunlight on the suicide rates based on the same model as in Table S2 Column (1) but at different levels of clustering the standard errors. In particular, the standard errors are clustered at the county level in Column (1) as the reference from the main analysis, the county + state-by-year level in Column (2), the county + year level in Column (3), and the state level in Column (4). The interpretations of the parameters are the same as Table S2. N = 444,861.
\*\*\* $p < 0.01$ , \*\* $p < 0.05$ , \* $p < 0.1$

**Table S5:** Robustness to Inclusion of Air Pollution

|  | (1) | (2) | (3) |
| --- | --- | --- | --- |
|  | Baseline | Add PM2.5 | Add lagged PM2.5 |
| $\beta_0 + \beta_{-1}$ | -0.197** | -0.196** | -0.189** |
| $se(\beta_0 + \beta_{-1})$ | (0.084) | (0.084) | (0.086) |
| $p(\beta_0 + \beta_{-1})$ | [0.020] | [0.020] | [0.028] |

*Notes:* This table presents the results with controlling for the monthly average PM2.5 concentrations levels. The sample period is 2000–2004. The interpretations of the parameters are the same as Table S2. Column (1) replicates the main analysis in Table S2 Column (1). Column (2) adds the PM2.5 concentrations in a given month, and Column (3) adds the lags in PM2.5 as sunlight, i.e., the values in given and past two months as well as the following month. The samples in Columns (1) and (2) are selected to be consistent to Column (3). $N = 27,009$ .
\*\*\* $p < 0.01$ , \*\* $p < 0.05$ , \* $p < 0.1$

**Table S6:** Estimated Effects of a set of Environmental Factors

|  | (1) | (2) | (3) | (4) | (5) |
| --- | --- | --- | --- | --- | --- |
| $\ln(\text{Sunlight})_0$ | -0.009<br>(0.019) | -0.035*<br>(0.020) | -0.023<br>(0.022) | -0.049**<br>(0.022) | |
| $\ln(\text{Sunlight})_{-1}$ | -0.063***<br>(0.020) | -0.080***<br>(0.021) | -0.074***<br>(0.024) | -0.085***<br>(0.024) | |
| $\ln(\text{Sunlight})_{-2}$ | 0.002<br>(0.020) | 0.002<br>(0.020) | -0.005<br>(0.022) | -0.003<br>(0.022) | |
| $\ln(\text{Sunlight})_1$ | -0.005<br>(0.022) | 0.002<br>(0.022) | -0.000<br>(0.023) | 0.010<br>(0.024) | |
| $\text{Temp}_0$ | | 0.008***<br>(0.001) | | 0.008***<br>(0.001) | 0.007***<br>(0.001) |
| $\text{Temp}_{-1}$ | | -0.002**<br>(0.001) | | -0.002**<br>(0.001) | -0.002***<br>(0.001) |
| $\text{Temp}_{-2}$ | | 0.000<br>(0.001) | | 0.000<br>(0.001) | 0.000<br>(0.001) |
| $\text{Temp}_1$ | | 0.001<br>(0.001) | | 0.001<br>(0.001) | 0.001<br>(0.001) |
| $\text{Precip}_0$ | | | -0.044<br>(0.032) | -0.043<br>(0.032) | -0.006<br>(0.029) |
| $\text{Precip}_{-1}$ | | | -0.029<br>(0.036) | -0.014<br>(0.036) | 0.041<br>(0.032) |
| $\text{Precip}_{-2}$ | | | -0.017<br>(0.033) | -0.013<br>(0.034) | -0.017<br>(0.030) |
| $\text{Precip}_1$ | | | 0.011<br>(0.032) | 0.019<br>(0.032) | 0.013<br>(0.030) |
| _cons | 1.677***<br>(0.405) | 1.926***<br>(0.411) | 1.939***<br>(0.459) | 2.088***<br>(0.461) | 0.875***<br>(0.029) |
| $\beta_0 + \beta_{-1}$ | -0.072 | -0.115 | -0.097 | -0.134 | |
| $\text{se}(\beta_0 + \beta_{-1})$ | 0.028 | 0.028 | 0.030 | 0.031 | |
| $p(\beta_0 + \beta_{-1})$ | 0.009 | 0.000 | 0.001 | 0.000 | |

Notes: Columns (1)–(4) present the effects of sunlight on suicides with various sets of other controls, whereas Column (5) presents the effects of temperature and precipitation on suicides without controlling for sunlight. The interpretations of the bottom parameters are the same as Table S2.
\*\*\* $p < 0.01$ , \*\* $p < 0.05$ , \* $p < 0.1$

**Table S7:** Estimated Effects of Sunlight on Temperature

|  | (1) | (2) | (3) | (4) | (5) | (6) |
| --- | --- | --- | --- | --- | --- | --- |
|  | Weighted |  |  | Unweighted |  |  |
| ln(Sunlight) |  |  | 2.633***<br>(0.177) |  |  | 1.479***<br>(0.079) |
| Precipitation |  | -2.319***<br>(0.236) | -0.543***<br>(0.193) |  | -2.182***<br>(0.060) | -1.173***<br>(0.072) |
| R<sup>2</sup> | .9727 | .9728 | .9731 | .9708 | .9709 | .9710 |
| $\Delta Temp$ (°C) | | | -1.328 | | | -.732 |
| $\Delta Temp$ (SD) | | | -0.144 | | | -0.074 |

*Notes:* This table presents the estimated effects of sunlight and precipitation as well as the county-by-month and state-by-year fixed effects on temperature. Columns (1) and (4) include only the fixed effects. Columns (1)–(3) are weighted by population as in the main analysis, whereas Columns (4)–(6) are not weighted. The last two rows indicate the effect of a 1-standard deviation (SD) decrease in sunlight on temperature in °C and SD, respectively.
\*\*\* $p < 0.01$ , \*\* $p < 0.05$ , \* $p < 0.1$

**Table S8:** Estimated Effects of Sunlight on Suicides based on Time-Series Analysis

|  | (1) | (2) | (3) | (4) |
| --- | --- | --- | --- | --- |
| $\ln(\text{Sunlight})_0$ | 0.050***<br>(0.016) | 0.041**<br>(0.016) | 0.044***<br>(0.016) | 0.044***<br>(0.016) |
| $\ln(\text{Sunlight})_{-1}$ | -0.016<br>(0.016) | -0.019<br>(0.016) | -0.019<br>(0.016) | -0.019<br>(0.016) |
| $\beta_0 + \beta_{-1}$ | 0.034 | 0.022 | 0.025 | 0.026 |
| $\text{se}(\beta_0 + \beta_{-1})$ | 0.013 | 0.013 | 0.014 | 0.013 |
| $p(\beta_0 + \beta_{-1})$ | 0.012 | 0.104 | 0.071 | 0.057 |
| Fixed effects | County | County +<br>year | County +<br>state-by-year | County-by-year |

*Notes:* This table shows the estimated effects of sunlight on suicides based on the time-series analysis that mimics the literature. In particular, we run the same model as in Table S2 Column (1) but with different fixed effects as specified in each column. The interpretations of the bottom parameters are the same as Table S2. N=444,864.
\*\*\* $p < 0.01$ , \*\* $p < 0.05$ , \* $p < 0.1$

**Table S9:** Effects of Sunlight on Depressive Language Searches from Google Trends

|  | (1) | (2) | (3) | (4) |
| --- | --- | --- | --- | --- |
| *Panel A: All keywords* |  |  |  |  |
| Effect | -8.464*** | -10.896*** | -5.860** | -5.549** |
|  | (1.038) | (2.608) | (2.817) | (2.306) |
| *p* | [0.000] | [0.000] | [0.039] | [0.017] |
| N | 4655 | 8796 | 8774 | 8774 |
| Mean of dep. var | 64.345 | 60.471 | 60.406 | 60.406 |
| Mean of sunlight | 16764.72 | 19018.18 | 19025.59 | 19025.59 |
| *Panel B: depression, depressed, depress* |  |  |  |  |
| Effect | -8.881*** | -12.557*** | -8.697 | -7.746 |
|  | (2.426) | (4.805) | (6.388) | (6.158) |
| *p* | [0.001] | [0.010] | [0.175] | [0.210] |
| N | 4655 | 8785 | 8763 | 8763 |
| Mean of dep. var | 60.509 | 53.439 | 53.413 | 53.413 |
| Mean of sunlight | 16764.72 | 19018.61 | 19026.03 | 19026.03 |
| *Panel C: suicide, suicidal* |  |  |  |  |
| Effect | -4.584*** | -7.545 | -5.507 | -5.287 |
|  | (1.193) | (4.632) | (5.278) | (5.006) |
| *p* | [0.000] | [0.105] | [0.298] | [0.292] |
| N | 4655 | 8697 | 8675 | 8675 |
| Mean of dep. var | 60.589 | 51.585 | 51.537 | 51.537 |
| Mean of sunlight | 16764.72 | 19024.99 | 19032.45 | 19032.45 |
| Region type | State | DMA | DMA | DMA |
| Fixed effects | State | DMA | DMA | DMA |
|  | + year | + year | + state × year | + state × |
|  | + month | + month | + state × | month |
|  |  |  | month | + state trend |

Notes: Each panel reports the effect of a one-log point increase in sunlight in given and previous months on the Google Trends index in the first row, the standard errors clustered at the region level in the second row, the *p*-value at the third row, the number of observations in the fourth row, the mean value of the dependent variable in the fifth row, and the mean value of sunlight (KJ/m<sup>2</sup>) in the last row. The dependent variable is scaled from 0 to 100 with respect to the highest point in each region (see Methods for data processing). Column (1) uses observations at the state-year-month level, and Columns (2)–(4) use observations at the DMA-year-month level.
\*\*\**p*<0.01, \*\**p*<0.05, \**p*<0.1

**Table S10:** Marginal Impacts of Weather vs. Climate

|  | (1) | (2) | (3) | (4) | (5) |
| --- | --- | --- | --- | --- | --- |
| Climate ($\beta$) | -0.078***<br>(0.017)<br>[0.000] | -0.070***<br>(0.018)<br>[0.000] | -0.081***<br>(0.018)<br>[0.000] | -0.078***<br>(0.017)<br>[0.000] | -0.076***<br>(0.017)<br>[0.000] |
| Weather ($\delta$) | -0.124***<br>(0.030)<br>[0.000] | -0.129***<br>(0.033)<br>[0.000] | -0.130***<br>(0.029)<br>[0.000] | -0.146***<br>(0.029)<br>[0.000] | -0.115***<br>(0.030)<br>[0.000] |
| Fixed effects | County +<br>month +<br>state × year | County +<br>month ×<br>year | County +<br>month +<br>year | County +<br>month +<br>year +<br>state-trend | County +<br>month +<br>county ×<br>year |

*Notes:* This table reports the estimated effects from Equation (5) in Methods. “Climate ( $\beta$ )” reports the $\beta$ ’s and “Weather ( $\delta$ )” reports the $\delta$ ’s from Equation (5). The standard errors, clustered at the county level, are reported in the parentheses, and the $p$ -values are reported in the square brackets. The sample size is 451,371.
\*\*\* $p < 0.01$ , \*\* $p < 0.05$ , \* $p < 0.1$

**Table S11:** Changes in sunlight required to achieve the equivalent impacts of other interventions

| Intervention | %change in suicide rate |  |  | Equivalent % change in sunlight |  |  | Equivalent SD change in sunlight |  |  |
| --- | --- | --- | --- | --- | --- | --- | --- | --- | --- |
|  | Estimate | Lower bound | Upper bound | Estimate | Upper bound | Lower bound | Estimate | Upper bound | Lower bound |
| 10-ug/m2 change in PM10, meta-analysis | 2 | 0 | 3 | -13.28 | 0.00 | -19.25 | -0.34 | 0.00 | -0.49 |
| 1C increase in monthly average temperature, US | 0.42 | 0.21 | 0.63 | -2.95 | -1.49 | -4.39 | -0.08 | -0.04 | -0.11 |
| State firearm regulations, US | -5.6 | -7.4 | -3.9 | 49.05 | 69.45 | 32.04 | 1.25 | 1.77 | 0.82 |
| National suicide prevention program, OECD countries | -6.62 | -11.31 | -1.93 | 60.31 | 123.92 | 14.77 | 1.54 | 3.16 | 0.38 |
| Celebrity suicide, JPN | 4.6 | 2.4 | 6.7 | -27.95 | -15.72 | -37.97 | -0.71 | -0.40 | -0.97 |
| 1% increase in suicide, EU | 0.49 | -0.04 | 1.02 | -3.43 | 0.29 | -7.01 | -0.09 | 0.01 | -0.18 |
| COVID-19, JPN | 16 | 11 | 21 | -68.03 | -54.34 | -77.61 | -1.73 | -1.38 | -1.98 |

*Notes:* This figure describes the amount of changes in sunlight in both percentage and standard deviation (SD) that is required to achieve the equivalent impacts of other interventions estimated in previous studies (25,24,36–40), “Intervention” describes the type of intervention, “Equivalent % change in suicide rate” describes the percentage change in suicide rate by the intervention, “Equivalent % change in sunlight” indicates the percentage change in sunlight that is required to achieve the same magnitude of impact as the intervention, and “Equivalent SD change in sunlight” describes the change in the standard deviation of sunlight that is required to achieve the same magnitude of the impact as the intervention. Each number under “Estimate” represents the mean impact, and those under “Lower bound” and “Upper bound” represent the 95% CI.

**Table S12:** Projected impacts of solar radiation management geoengineering on suicides

| Temperature limit | Sunlight | Temperature | Net |
| --- | --- | --- | --- |
| *Panel A: 0%* |  |  |  |
| 1.5 °C | 2.23 (1.26, 3.18) | -5.75 (-10.69, -1.23) | -3.52 (-7.72, 0.781) |
| 2 °C | 1.46 (0.83, 2.08) | -3.77 (-7.00, -0.81) | -2.31 (-5.06, 0.509) |
| *Panel B: 1%* |  |  |  |
| 1.5 °C | 1.78 (1.01, 2.55) | -4.61 (-8.56, -0.99) | -2.82 (-6.19, 0.623) |
| 2 °C | 1.02 (0.58, 1.45) | -2.63 (-4.89, -0.56) | -1.61 (-3.53, 0.354) |
| *Panel C: 3%* |  |  |  |
| 1.5 °C | 1.22 (0.70, 1.75) | -3.17 (-5.89, 0.68) | -1.94 (-4.26, 0.426) |
| 2 °C | 0.47 (0.27, 0.67) | -1.22 (-2.26, -0.26) | -0.746 (-1.63, 0.163) |
| *Panel D: 5%* |  |  |  |
| 1.5 °C | 0.91 (0.52, 1.30) | -2.35 (-4.37, -0.50) | -1.44 (-3.16, 0.316) |
| 2 °C | 0.17 (0.10, 0.24) | -0.44 (-0.81, -0.09) | -0.267 (-0.585, 0.058) |

*Notes:* This table summarizes the cumulative additional suicides due to the negative radiative forcing (in the second column) and averted suicides due to temperature fall (in the third row) to meet the goal of keeping global temperature rise below 1.5 °C in the first row and 2 °C in second row of each panel. The last column represents the net effect of sunlight and temperature effects estimated from the simulation (see Methods). The numbers in the parentheses represent the 95% CI. All numbers are in thousands of deaths in 2100. Each panel title refers to annual emissions reduction rate between 2031 and 2100.

**Figure S1:**
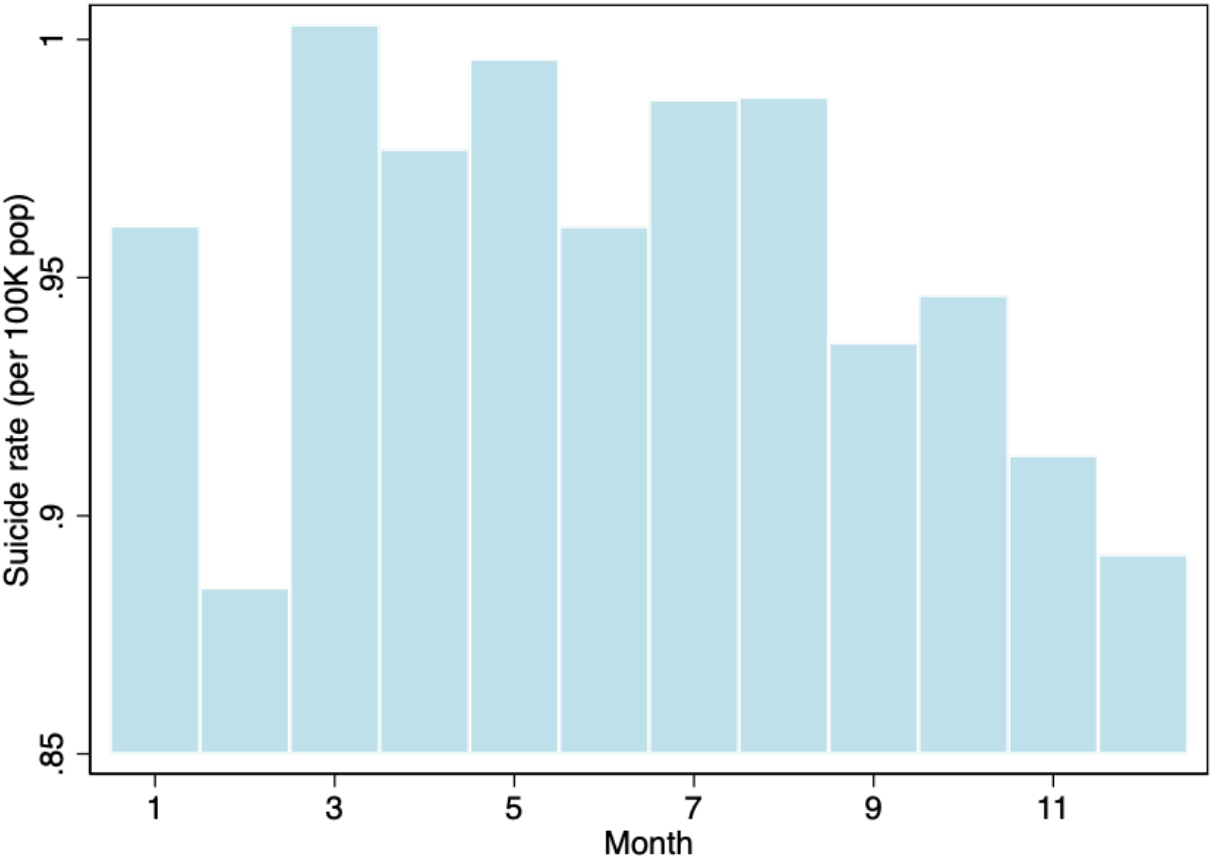
Seasonality Trends in the Suicide Rate *Notes*: This figure shows the monthly average suicide rates based on the data for the analysis.

**Figure S2:**
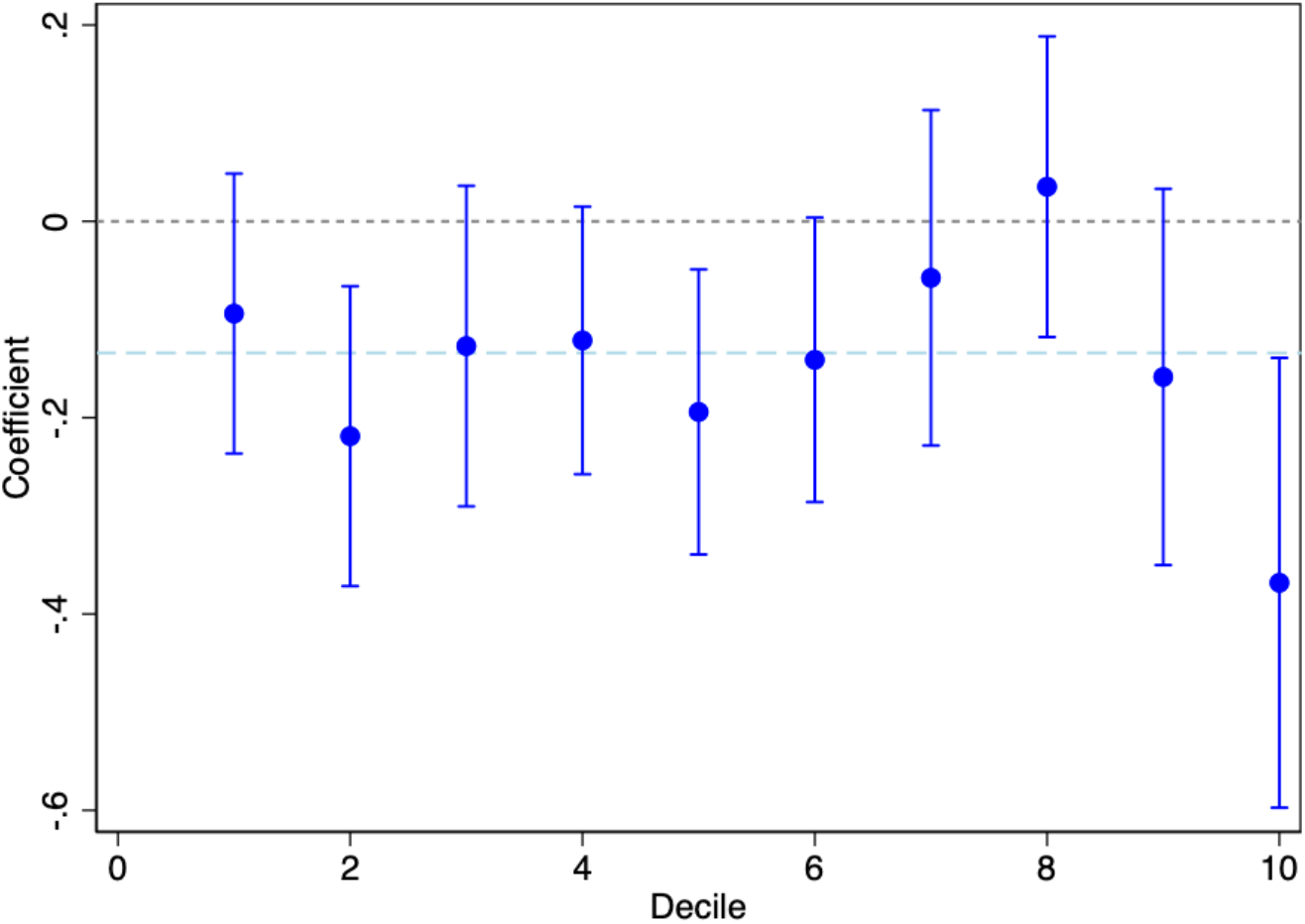
The Effects of Sunlight on the Suicide Rate by Decile *Notes*: This figure plots the cumulative effect of sunlight in given and previous months, as given by *β*_0_ + *β*_−1_ and its 95% confidence interval in each decile bin of the average amount of sunlight at the county level. The regression additionally controls for the average temperature and precipitation in given and previous months as well as the county-by-month and state-by-year fixed effects. The regression is weighted by county population. The standard errors are clustered at the county level. The blue dashed line indicates the estimated effect from the main analysis for the reference purpose.

**Figure S3:**
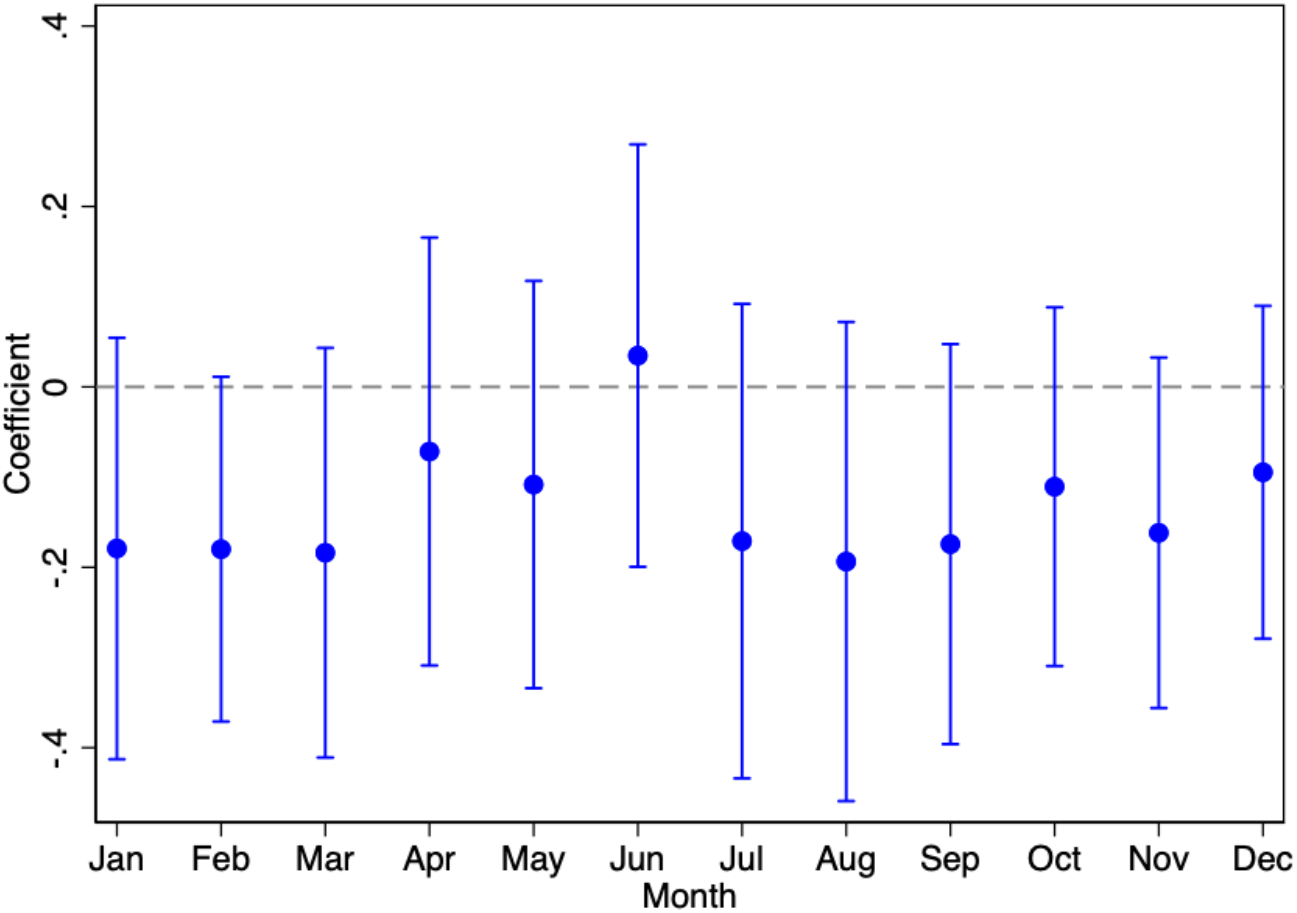
The Effects of Sunlight on the Suicide Rate by Month *Notes*: This figure plots the cumulative effect of sunlight in given and previous months, as given by *β*_0_ + *β*_−1_ and its 95% confidence interval in each month. The regression additionally controls for the average temperature and precipitation in given and previous months as well as the county-by-month and state-by-year fixed effects. The regression is weighted by county population. The standard errors are clustered at the county level.

**Figure S4:**
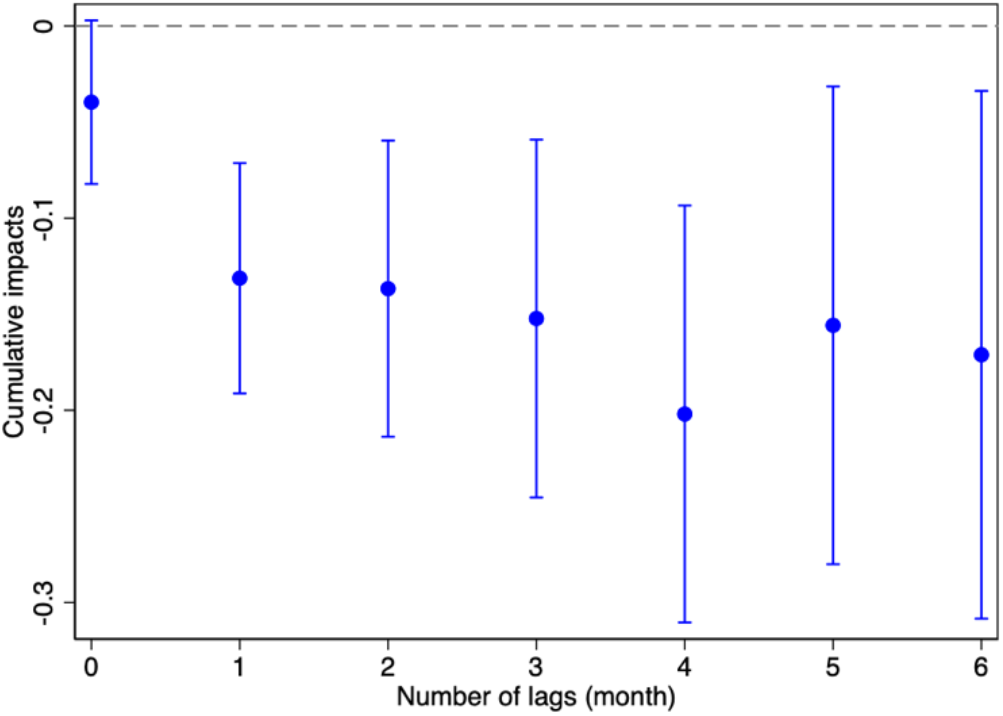
Robustness to Extensive Temporal Displacements *Notes*: This figure plots the cumulative impacts on the suicide rate of the log of sunlight exposure in given and previous months. Each value reports 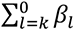 and the associated 95% CI from a separate regression for each *l* ∈ [0,6], as we iteratively add an additional month in the past. All regressions additionally control for the monthly average temperature and precipitation with the same set of lags, the log of sunlight, temperature, and precipitation in the following month, and county-by-month and state-by-year fixed effects.

**Figure S5:**
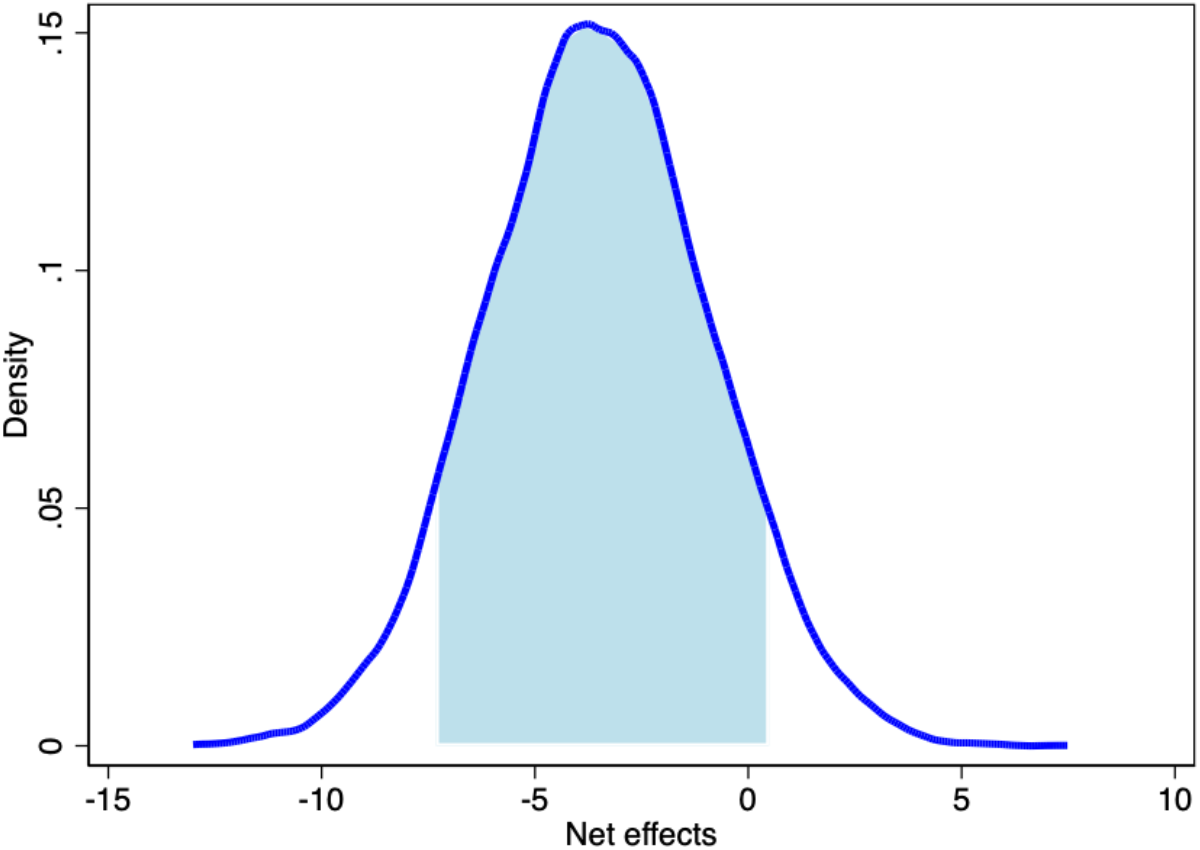
Projected cumulative net impacts of solar radiation management on suicides in 2030–2100 *Notes*: This figure plots the cumulative net impacts of excess suicides due to reduced sunlight and averted suicides due to lower temperature to meet the goal of keeping the global temperature rise below 1.5 °C in 2030–2100. The values are based on the simulation of 10,000 replications based on the parameters described in Methods. The shaded area represents the 95% of the distribution.

